# Quantitative Assessment of Climate Change Effects on Global FoodPrices: Evidence from the North Atlantic Oscillation Index

**DOI:** 10.64898/2026.02.26.26347157

**Authors:** Kaies Ncibi

## Abstract

Food costs are more significantly impacted by climate change as countries grow. It is well known that climate change has an impact on the productivity of most agricultural goods, but it is unclear how specifically it will affect food costs. The present research explores how the North Atlantic Oscillation (NAO) index, a widely used climate indicator, affects food prices around the world. This is achieved by applying a robust bivariate Hurst exponent (robust bHe). The research creates a color map of this coefficient using a window-sliding technique over various intervals of time, displaying an illustration that changes overtime.

Additionally, the NAO index and global food prices are examined for causal connections using variable-lag transfer entropy using a window-sliding technique. The results show that notable rises in a number of international food prices for long as well as short periods are associated with significant increases in the NAO index. Furthermore, the causative function of the NAO index in influencing global food costs is confirmed by variable-lag transfer entropy.

Is highly recommended as it directly connects the research to actionable outcomes for policymakers and the overarching goal of sustainability and food security. This study provides the first direct evidence of a robust, long-range cross-correlation and causal link between the North Atlantic Oscillation (NAO) index and key global food prices. It introduces a novel, robust methodological framework to visualize this time-varying relationship, offering a critical tool for policymakers and forecasting models.

## I. Introduction

The stability of global food systems is a fundamental prerequisite for achieving sustainable development goals, particularly those related to zero hunger, climate action, and responsible consumption. However, this stability is increasingly undermined by the multifaceted impacts of climate change, which introduce significant volatility into agricultural production and, consequently, international markets. While the negative biophysical effects of a changing climate on crop yields are increasingly well-documented, a critical and less understood nexus lies in the quantitative transmission of these climate shocks into global food price dynamics. Understanding these transmission channels is not merely an academic exercise; it is a pressing necessity for designing effective early warning systems, evaluating climate adaptation policies, and formulating market mechanisms that can ensure food security in an era of environmental uncertainty.

The existing economic and climatological literature has made substantial progress in linking large-scale climate phenomena to agricultural outcomes. A significant body of work has established the El Niño-Southern Oscillation (ENSO) as a key driver of price volatility for specific commodities like maize, soybeans, and rice, demonstrating its capacity to disrupt weather patterns across critical growing regions [40, 41, 65]. In a parallel research stream, the North Atlantic Oscillation (NAO)a major climate mode governing atmospheric mass variability between the subtropical Atlantic and the Arctic has been extensively studied for its direct, biophysical impact on winter weather in Europe, North America, and Asia, with demonstrated consequences for regional yields of cereals, soybeans, and other staples [44, 60, 72].

Despite this progress, a significant conceptual and empirical gap persists between these two research streams. The current paradigm largely follows a two-step logic: climate affects yields, and yields, in turn, influence prices. What remains inadequately explored is the potential for climate indices, specifically the NAO, to act as systematic and direct drivers of global food price dynamics, operating through complex, long-memory market mechanisms that may extend beyond immediate yield shocks. This gap is critical; the absence of a direct, quantified link between the NAO and global prices limits the ability of policymakers, commodity traders, and risk managers to incorporate this predictable climate indicator into strategic planning. Without such tools, efforts to build resilient food systems and implement proactive, rather than reactive, food security strategies are inherently constrained.

This study seeks to bridge this gap by moving beyond traditional regional yield analysis. We posit that the NAO index exerts a significant and measurable influence on global food prices that can be captured through sophisticated econometric techniques designed for long-range, power-law dependencies. Our research shifts the focus from regional production shocks to global market signals, directly quantifying the dynamic, long-memory connection between the NAO index and a basket of critical global food commodities: corn, soybean, wheat, and oats.

From a sustainability and management accounting perspective, this investigation is paramount. Clarifying this relationship provides a quantitative basis for several critical applications:

Informing Sustainable Pricing Mechanisms: By understanding how climate oscillations drive price volatility, more robust financial instruments and insurance products can be designed to hedge risk for farmers and vulnerable economies. Policy Evaluation: The findings offer a new metric for assessing the economic efficacy of climate adaptation policies, measuring their success in buffering global markets from climate-driven price shocks. Strategic Forecasting: Integrating the NAO into forecasting models enhances the predictive power of early warning systems, allowing for more timely interventions to protect food-insecure populations and stabilize markets.

To this end, the primary objectives of this research are: To empirically investigate the existence and nature of long-range, power-law cross-correlations between the NAO index and global prices of corn, soybean, wheat, and oats, employing a novel robust bivariate Hurst exponent.To determine the direction of causality and information flow between this climate indicator and food market volatility using a variable-lag transfer entropy approach, establishing whether the NAO acts as an exogenous driver.To demonstrate, through a sliding-window framework, how this time-varying relationship can be visualized and operationalized, providing a practical tool for integration into forecasting and risk management platforms.

The study is guided by the following research questions: Does a statistically significant, long-range cross-correlation exist between the NAO index and key global food prices, and how does this relationship evolve over time? Can the NAO index be established as a causal driver of price volatility in these markets, with a unidirectional flow of information? What are the tangible implications of this relationship for designing climate-resilient food policies, sustainable market mechanisms, and enhanced forecasting tools for global food security?

By answering these questions, this research aims to provide a missing link in the climate-economy nexus, offering empirical evidence and a novel methodological framework to support more informed and sustainable management of the global food system.

## II. Litterarure review

Significant issues have emerged due to the global COVID-19 pandemic, particularly in the realms of poverty and undernourishment. With consistently elevated rates of global malnutrition, research suggests that by the end of 2022, 75 million individuals may descend into severe poverty [25, 82, 86]. In light of this, this study seeks to illuminate one particular aspect that composes the intricate relationship between climate fluctuations and global food prices. Prior to that epidemic, 828 million individuals were facing starvation in the year 2021, up an astounding 150 million from 2019 and about 46 million over 2020 [1, 31]. Countries that have imposed limits on food trading in an effort to stabilize their own supplies and reduce price volatility have made the world’s hunger problem worse [53,58]. Worldwide food production has also been affected by changes in precipitation patterns, worldwide temperature changes, and the increased frequency of catastrophic storms, in addition to international trade mechanisms [17, 28]. World agricultural output has decreased by 21% from 1961 due to climate change, endangering economic growth, especially in developing countries [86]. This research aims to clarify the unique influence of the North Atlantic Oscillation (NAO) index on the worldwide cost of food in such a complicated worldwide setting. Worldwide crop output is significantly impacted by the NAO index, which measures the movement of atmospheric mass across the subtropical Atlantic and the Arctic or subarctic areas [45, 67]. Additionally, it has a significant impact on how winters are shaped in Asia, Europe, as well as the US, which in turn affects agricultural output [74, 83, 85].

The present study finds an absence in the investigation of the NAO index’s direct influence on global food prices, despite the fact that numerous investigations have examined its effect on agricultural productivity. Nonetheless, studies have examined the relationship amongst particular food items and various climate indicators, like the El Niño and La Niña indicators. For example, [65] looks at the way El Niño as well La Niña events affect maize and soybeans prices, showing that El Niño causes more volatility over the Spring-Summer period also have varying impacts on soybean prices depending on the period of time. Parallel to this, [81] looks at monthly grain futures prices across different areas to show price movements following El Niño and La Niña occurrences. [40] highlights the persistent short- and long-term effects of La Niña occurrences by establishing an advantageous relationship between grain export prices and these occurrences. [80] examines the fluctuations of the price ratio between fishmeal and soybean flour, showing that the bi-monthly multivariate El Niño Southern Oscillation (ENSO) has a major effect on the pricing ratios until as long as twelve months after ENSO events. Furthermore, [41] observes a negative correlation with La Niña shocks but a positive correlation with rice prices and the El Niño index.

Many investigations have examined the influence of the NAO index on food prices in different areas and nations, but there is currently no direct investigation into the possible relationship involving the index and global food prices. Whereas [72] offers proof connecting the NAO index with soybean output in Italy, [44] demonstrates the impact of the index on soybean production in Australia plus Europe. Additionally, [3] illustrates the worldwide aggregation variation in grain, soybean, and corn production, which is ascribed to the NAO index, the Indian Ocean Dipole, the ENSO index, and tropical Atlantic variation, correspondingly. Additionally, [60] shows an important connection among the variation of soybean output among countries and large-scale fluctuations in the NAO index. The association involving the NAO index and corn, rice, wheat, barley, oats, and potatoes is examined through research such as [93] and [12], which show that the connections differ depending on the crop kind and changing seasons.

While extant literature has established broad correlations between climate phenomena and agricultural yields, this study provides a significant methodological and empirical advancement. It moves beyond regional or single-commodity analyses to directly quantify the dynamic, long-memory connection between a key climate indicator the North Atlantic Oscillation (NAO) index and a basket of critical global food prices. Via integrating a robust bivariate Hurst exponent and utilizing variable lag transfer entropy within a sliding-window methodology, we distinctly record and illustrate the dynamic, multi-scale cross-correlation and causal information flow. This technique absolutely not only validates a statistically significant power-law relation but also identifies the NAO index as a direct causal influencer of global price volatility, providing a more accurate instrument for forecasting and risk management than previously offered.

Table 1 presents a concise examination of the relationship between various climate change indices and food prices, along with an outline of the methodology employed in these studies.

**Table 1:**
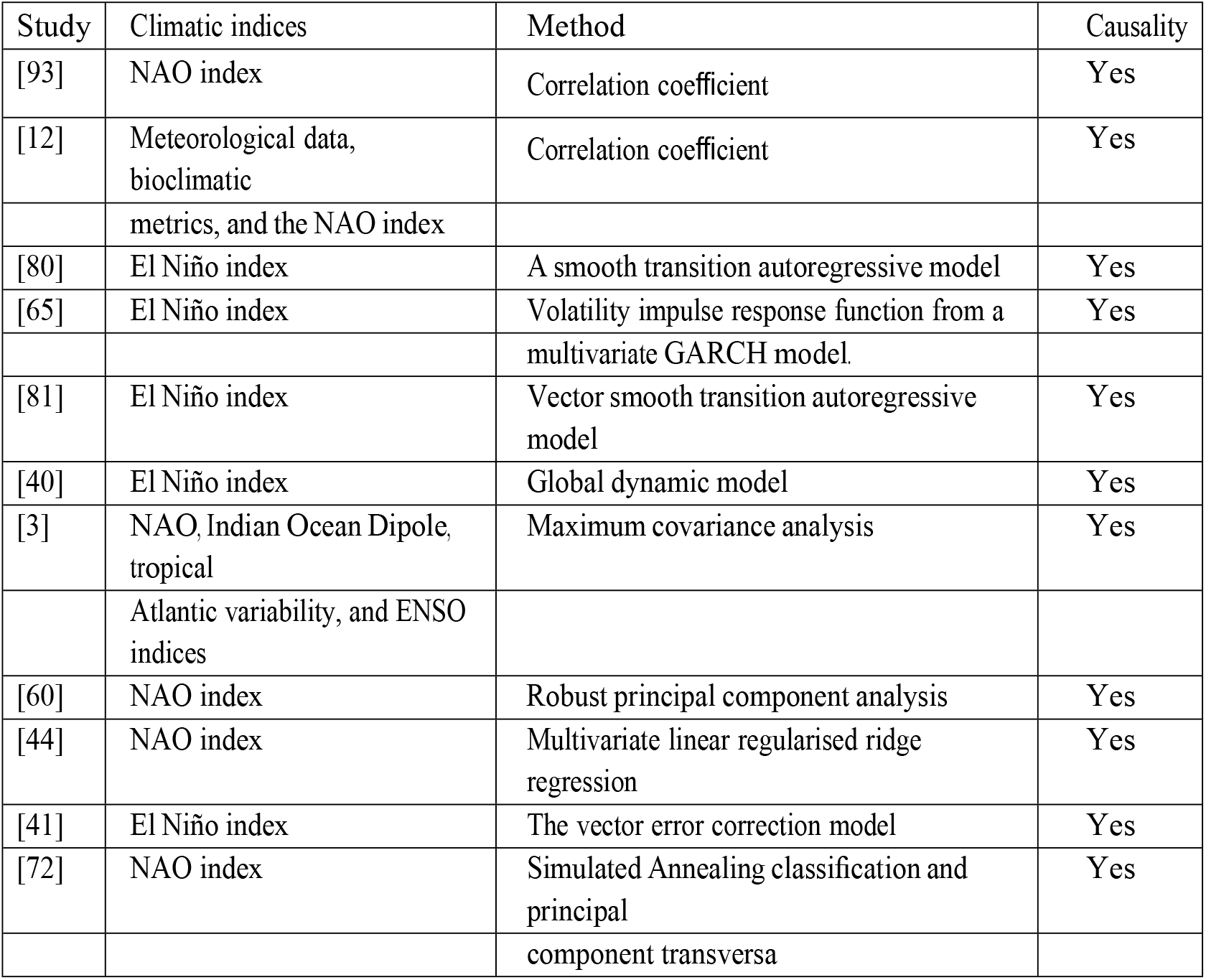
An overview of earlier studies.

This document outlines its structure as follows: The literature review is addressed in Section 1. We outline the methodologies utilized in this study in Section 2. Section 3 includes descriptive statistics alongside a description of the sample data. Section 4 offers the robust bivariate Hurst exponent findings. Following this, Section 5 reveals the outcomes of the variable-lag transfer entropy causality assessment. Section 6 examines the implications, both theoretical and practical. In conclusion, Section 7 summarizes our findings. A significant volume of research has established the detrimental impacts of climate change on agricultural yield, forming a distinct theoretical connection to food security [17, 28, 86]. Within this domain, numerous investigations have effectively correlated particular climate indices, notably the El Niño-Southern Oscillation (ENSO), with the price fluctuations of specific commodities such as maize, soybeans, and rice [40, 41, 65, 80, 81]. Likewise, studies concerning the North Atlantic Oscillation (NAO) have primarily concentrated on its immediate biophysical effects on crop productivity and regional outputs in Europe, North America, and Australia [44, 60, 72, 93]. Nevertheless, a significant void remains between these two research pathways. While existing literature firmly establishes that climate influences yields and that ENSO affects prices, it does not provide a direct examination of the NAO’s function as a systematic catalyst for global food price trends.

Prior work has either examined the NAO’s influence on regional agricultural outputs or correlated other climate indices with markets, leaving the specific, quantitative linkage from the NAO to global price volatility unexplored. This study addresses this gap by moving beyond regional yield analysis to directly investigate the long-range cross-correlation and causal information flow between the NAO index and a basket of key global food commodities, employing a novel robust methodology capable of capturing this complex, time-varying relationship.

## III. Methodology

The present research offers a strong analytical approach to analyze the dynamic, time-varying relationship between the NAO index and global food prices. Data preparation, correlation analysis, and causation testing are the three main steps in the methodology’s progression.

### 3.1. Key Analytical Techniques

#### 3.1.1. Robust Bivariate Hurst Exponent (bHe) for Long-Range Correlation

Common correlation measures (e.g., Pearson, Spearman) reflect short-term, linear correlations. Researchers use the Robust Bivariate Hurst Exponent (bHe) [66] to identify long-term, durable connections that could possibly be non-linear and function on several time levels. This technique has been developed to be resistant to outliers and is based on detrended cross-correlation analysis [41].

The power-law scaling of cross-correlations between two series, x (NAO index) and Y (food price returns), is quantified by the robust bHe, represented by *H*_*X,Y*_. The calculation entails dividing the series into separate windows, computing the scaling of the q^th^-order cross-correlation function window: 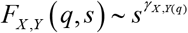 with window size s, and decreasing inside every window:

Assuming q=2, we estimate *H*_*X,Y*_ to be the scaling exponent *γ*_*X,Y*_ (2). A value of *H*_*X,Y*_ = 0.5 denotes no cross-correlation, *H*_*X,Y*_ > 0.5 exhibits persistent positive cross-correlation, while *H*_*X,Y*_ < 0.5 indicates anti-persistence [66]. Applying critical values obtained from a Monte Carlo simulation with 10,000 iterations, we compare the alternative hypothesis *H*_1_: *H*_*X*_ + *H*_*Y*_ > 1 versus the null hypothesis *H*_0_: *H*_*X*_ + *H*_*Y*_ = 1 (no cross-correlation) [66].

Let X (t) and Y (t) be two time series with zero mean, exhibiting long-range power-law autocorrelation:

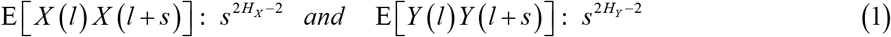

Where *H*_*X*_ *and H*_*Y*_ represent the Hurst exponents, falling within the range of [0.5, 1[. The power-law cross-correlations are defined as:

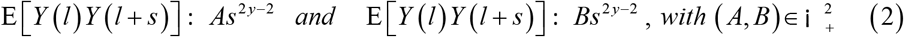

The phases for the robust bHe assessment are outlined below:

- **Profile construction:** We divide each time series into non-overlapping boxes of size *s*, where *s* is defined as *N* / *s*,and *l*_*v*_ = (*v* −1) *s*. The profiles in the v-th box are constructed as follows:

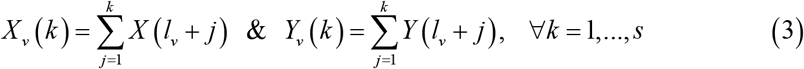

Trend estimation: we estimate the local trends of *X*_*v*_ (*k*) *and Y*_*v*_ (*k*), denoted by 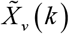 *and* 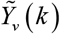, respectively.
- **Cross-correlation calculation:** The cross-correlation for each box is computed using the following equation:

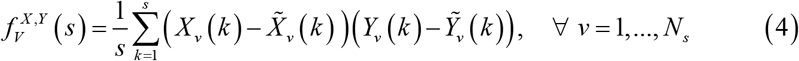
- **q-th order cross-correlation function:** The q-th order cross-correlation function is determined as follows:

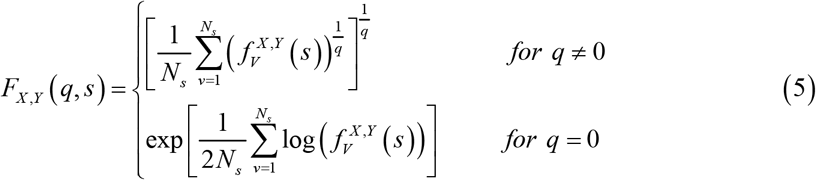
- **Scaling relation:** For sufficiently large ^*s*^, we expect a scaling relation as follows:

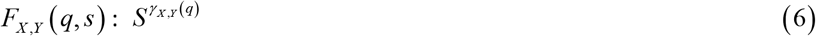

Where the Hurst exponent *γ*_*X,Y*_ (*q*) characterizes the long-range cross-correlation properties of processes X and Y. Avalue of *γ*_*X,Y*_ (2) around 0.5 indicates no cross-correlation, while *γ*_*X,Y*_ (2) > 0.5 suggests a positive power-law cross-correlation, and *γ*_*X,Y*_ (2) < 0.5 indicates anti-correlation.

#### 3.1.2. Sliding-Window Implementation for Time-Varying Analysis

To observe the way the long-range correlation develops, we construct the robust bHe within a sliding-window architecture. With a window of dimension h and a time range n (e.g., 50, 100, 150, 200 days), we compute *H*_*X,Y*_ (*T, n*) for every window beginning at time T. This provides a matrix of bHe values over time Tand size n, that may be displayed as a color map (see Results, Figures 6-9), highlighting times where the NAO-price link strengthens or decreases. The time-varying cross-correlation can be summarized by summing across scales:

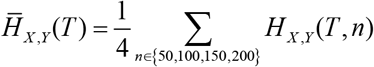

Given every window beginning at time T : 1 ≤ *T* ≤ *N* − *h* :

- Generate sub-series : 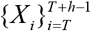 *and* 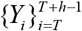.
- Find the local bHe, *H*_*X,Y*_ (*T, n*) assuming a fixed scale n using the method in Section 3.1.
- Cycle for each scales n and every window beginning times T.

#### 3.1.3. Variable-Lag Transfer Entropy (VLTE) for Causality

Causality is not implied by connection. We use Variable-Lag Transfer Entropy (VLTE) to determine the direction of information flow [2]. Transfer entropy quantifies the decrease in uncertainty regarding the future outcomes of a variable Y (food price returns) knowing the historical data of another variable X (NAO index), beyond what is included in Y’s original history [29, 86]. The VLTE extension is significant since it enables flexible, changeable intervals among cause and effect, which is realistic for climate-economic systems [2].

We calculate the VLTE via *X* → *Y* as well as *Y* → *X*.

A VLTE ratio larger than 1, *VLTE* 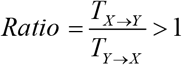, shows that X has a unidirectional causality impact on Y [2]. To establish consistency and track changes in causality power as time goes on, we conduct this test with identical sliding-window variables as the bHe study.

This approach offers a thorough evaluation of how the NAO index influences volatility in international food markets while taking into account intricate, time-varying dynamics by combining two reliable, complementary approaches: one of them for measuring persistent correlation [66] and a second to determine produced information flow [2, 29].

### 3.2. Descriptive statistical data

The time series under consideration comprise the global prices of corn, soybean, oats, and wheat (in U.S. dollars per bushel), received via the Bloomberg Terminal, and the North Atlantic Oscillation (NAO) index, gathered by the NOAA Climate Forecast Unit. The data covers the period from January 6, 2020, to May 18, 2022, yielding a single-day frequency with 600 observations. 1 16 This specific timeframe was strategically selected to capture a period of exceptional and interconnected volatility. It encompasses the onset and persistent effects of the COVID-19 pandemic, which induced unprecedented disruptions to global supply chains, agricultural labor markets, and trade policies, thereby exerting significant pressure on food prices. Concurrently, this period featured pronounced climate variability, allowing for the analysis of the NAO’s influence against a backdrop of broader environmental and economic instability. By focusing on this turbulent epoch, the study is positioned to investigate the climate-price nexus under stress conditions, offering insights that are critical for building resilience against compound crises a key priority for sustainable development and food security management. We investigate the fluctuations of each time series, with the exception of the NAO index, by examining their returns, which are determined by *x*_*t* +1_ − *x*_*t*_. Figure 1 offers a graphic illustration of the studied time series, with red points signifying outlier observations.

**Figure 1.**
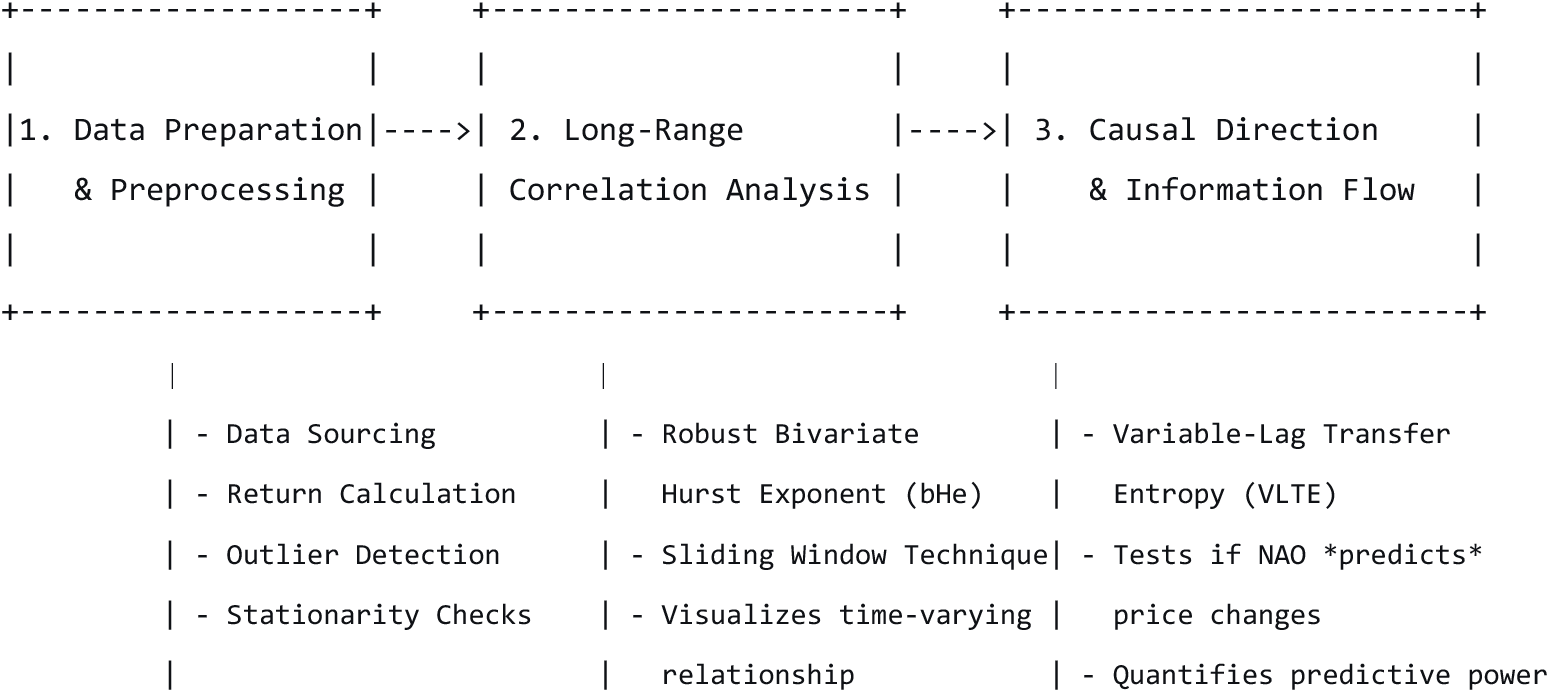
Analytical Framework for Assessing NAO-Food Price Dynamics

### 3.3. Data Sources and Variable Construction

This study analyzes the dynamic relationship between a key climate indicator and globalagricultural commodity markets. The dataset consists of:

- Climate Variable: The daily North Atlantic Oscillation (NAO) index. Data is publicly sourced from the National Oceanic and Atmospheric Administration (NOAA) Climate Prediction Center.
- Economic Variables: The daily closing prices (in U.S. dollars per bushel) for four critical agricultural commodities: corn, soybean, wheat, and oats. Price data is collected from the Bloomberg Terminal.

The data covers the time from January 6, 2020, to May 18, 2022, providing a total of T = 600 daily observations for each series. This timeframe was deliberately chosen to capture a period of compounded global volatility, including the supply chain disruptions and trade policy shifts of the COVID-19 pandemic, alongside significant climate variability. Analyzing this period allows for the investigation of the climate-price nexus under stress conditions, which is crucial for developing robust food security risk models. To model price dynamics and ensure stationarity for econometric analysis, the non-stationary price series are transformed into daily logarithmic returns. For a given commodity price *P*_*t*_, the daily return *R*_*t*_ is calculated as: *R*_*t*_ = ln(*P*_*t*_) − ln(*P*_*t* −1_).

This transformation reflects relative price changes and is the standard input for financial and economic volatility analysis. The NAO index, being a stationary climate anomaly index, is analyzed in its original form.

### 3.4. Data Cleaning and Preprocessing

A sophisticated multistage data cleaning and preparation procedure was put in place before analysis to guarantee temporal alignment, consistency, and robustness. The process tackled issues that arise when combining financial market data with daily climatic data.

## IV. Results

### 4.1. Descriptive Statistics and Preliminary Tests

Figure 2 presents a graphical representation of the 5 primary time sequences, featuring anomalous data points marked. The foundational statistical characteristics of the NAO index and the commodity return series are outlined in Table 2 (Panel A).

**Table 2(Panel A):** Descriptive Statistics of the Original Series.

| Series | Mean | Std. Dev. | EVT Outliers (%) | Breitung Test (p-value) | Robust JB Test (p-value) |
| --- | --- | --- | --- | --- | --- |
| NAO Index | -1.839 | 110.543 | 47 (1.958%) | 0.005 (0.001)*** | 98.554 (0.000)*** |
| Corn Returns | 0.0001 | 0.084 | 79 (3.291%) | 0.0002 (0.001)*** | 631,128.2 (0.000)*** |
| Soybean Returns | 0.001 | 0.160 | 73 (3.041%) | 0.0001 (0.001)*** | 11,679.66 (0.000)*** |
| Wheat Returns | 0.001 | 0.127 | 50 (2.083%) | 0.0002 (0.001)*** | 376,399.5 (0.000)*** |
| Oats Returns | 0.001 | 0.081 | 84 (3.500%) | 0.0001 (0.001)*** | 39,656.19 (0.000)*** |

\*Note: \*\*\* indicates significance at the 1% level.\*

**Figure 2:**
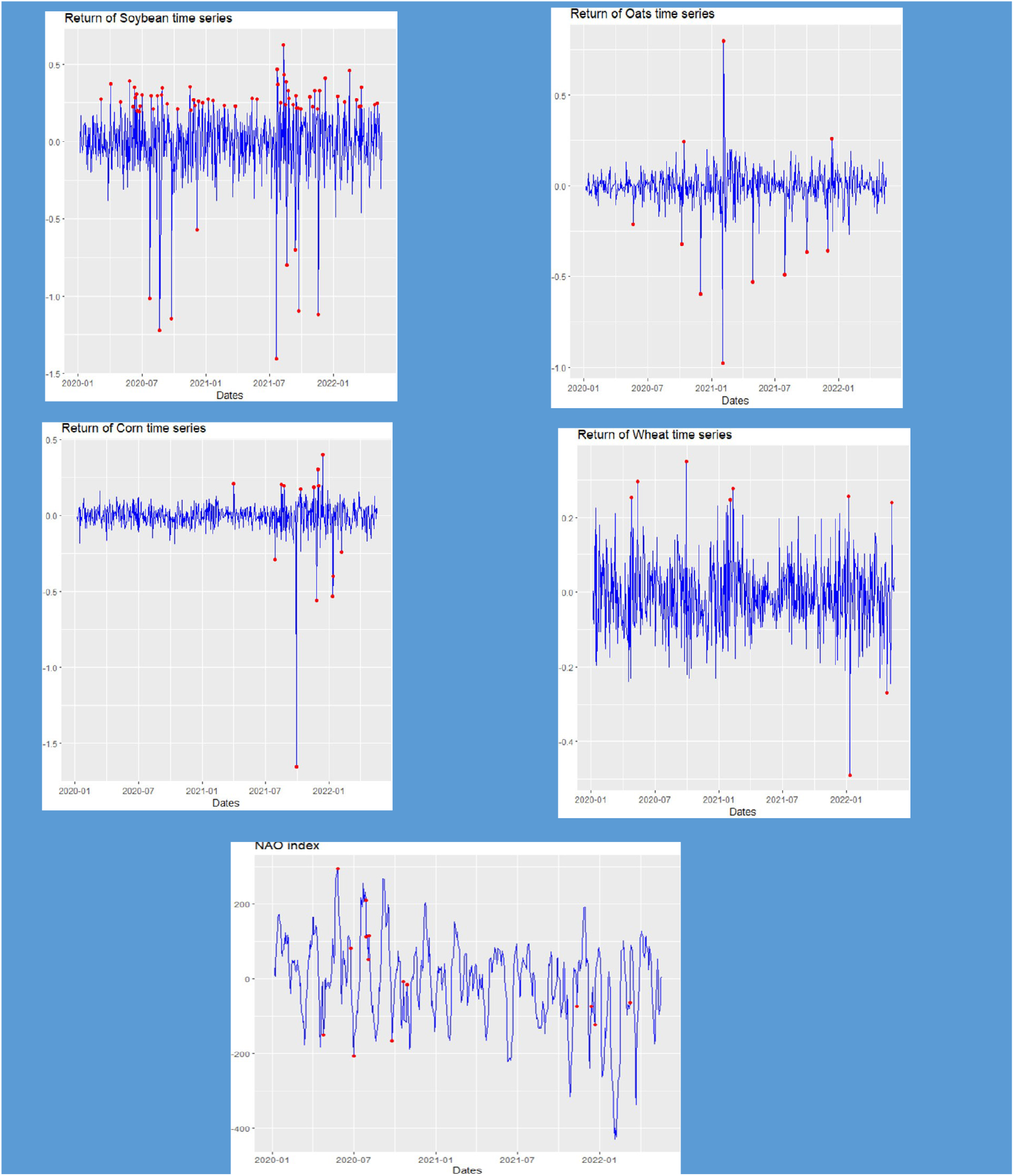
Explored time series

The main takeaways from Table 2 (Panel A) reveal the considerable fluctuations of the NAO index (Std. Dev. = 110.543) in comparison to the return series. The Extreme Value Theory (EVT) assessment affirms the existence of anomalies in all series. Most importantly, the resilient Breitung test dismisses the null hypothesis of a unit root for all series (p-values < 0.01), validating stationarity. The sturdy Jarque-Bera test also rejects the assumption of normality for each series (p-values < 0.01), which supports the application of non-parametric correlation methods.

Taking the non-standard distribution of the return series, Spearman’s rank correlation coefficient (ρρ) is utilized to evaluate initial bivariate relationships. The findings, presented in Table 3 (Panel B), show no notable linear correlation between the NAO index and any of the commodity returns at standard significance levels, underlining the importance of our advanced long-range and non-l

**Table 3(Panel B):** Stationarity, Normality, and Correlation Tests.

| Series | ADF Test (p-value) | JB Test (p-value) | $\rho\rho$-Spearman (p-value) |
| --- | --- | --- | --- |
| NAO Index | -5.7277 (0.01)*** | 1.6824 (0.431) | -- |
| Corn Returns | -13.243 (0.01)*** | 631,128 (0.000)*** | -0.009 (0.823) |
| Soybean Returns | -12.759 (0.01)*** | 11,680 (0.000)*** | -0.043 (0.286) |
| Wheat Returns | -14.019 (0.01)*** | 376,400 (0.000)*** | -0.026 (0.516) |
| Oats Returns | -14.243 (0.01)*** | 39,656 (0.000)*** | 0.045 (0.269) |

\*Note: \*\*\* indicates significance at the 1% level.\*

In order to explore the intricate connections between climatic trends and food costs, this research utilizes an innovative, multi-step investigative approach. The framework is crafted to withstand anomalies and adept at identifying long-term, non-linear relationships that conventional connection metrics may overlook. The entire procedure is illustrated in Figure 1 below.

As indicated by the ADF testing outcomes, all time series are stationary, as shown in Table 3 (Panel B), since all p-values are less than 0.05. Nevertheless the p-value is greater than 0.05, the JB test findings indicate that the NAO index is normally distributed. On the other hand, because the corresponding p-values of the JB test are less than 0.05, none of the additional time series are normally distributed. Our team used the Spearman coefficient to calculate the association among the NAO index and every examined worldwide food price because the outcomes on such prices do not follow a normal distribution.

Table 2 displays descriptive data for the time series under study. Different patterns amongst the time series become apparent upon close inspection of the results shown in Table 2 (Panel A). In particular, the NAO index is notable since it has a negative mean of −1.839, which is less than zero. A significant variance, represented by a standard deviation of 110.543, is also present. This highlights the significant fluctuation linked to the NAO index. On the other hand, the mean values for the remaining time series are identical to 0.001 for soybean, wheat, and oats, and 0.0001 for corn. Variances of 0.084, 0.16, 0.127, and 0.081 accompany this. In contrary to the more volatile character of the NAO index, such findings indicate an increase in mean returns across various products.

The final column of Table 3 displays the Spearman coefficient data, which show no discernible relationship among the NAO index and the other time series under study. As suggested, the robust bHe requires fractional Gaussian noise (fGn) processes that are tainted with additive outlier occurrences. We start by locating and eliminating outliers within the time series under study in order to calculate this factor. Next we determine the time series’ normality, stationarity, and stochasticity in the absence of outliers. The foundation for calculating the robust bHe is laid when the refined time series exhibits certain traits, which we regard as suggestive of a fGn system.

We use the extreme value theory (EVT) experiment, which was introduced by [46], to find outlier data in the time series under investigation. Table 2 (Panel A) displays the test results, which show the proportion of outliers in each time series. We use the unit root test based on Breitung’s variance ratio [11] to address the augmented Dickey-Fuller (ADF) and Kwiatkowski-Phillips-Schmidt-Shin (KPSS) tests’ sensitivity to the impact of outliers [32, 63]. Furthermore, we use the robust Jarque-Bera (robust JB) test [34] to evaluate the normality distribution of the time series under study. Table 2 (Panel A) provides specifics on the outcomes of these tests. According to the results of the outlier detection examination, there are 3.5%, 3.291%, 3.041%, 2.083%, and 1.958% outliers in the NAO index, the income of the oat, corn, soybean, and wheat time series, consecutively. Additionally, as all p-values are less than 1%, the robust stationarity test rejects the starting hypothesis of non-stationarity in support of the alternative hypothesis of stationarity. As a result, the time series under study are considered stationary. The robust Jarque-Bera (JB) test confirms that the distribution of the time series under study is not normal. The associated p-values (in parenthesis) are less than 1%, which means that the initial hypothesis of normality is rejected.

Table 5 displays the results of the stationarity and normality tests for the time series under study that do not contain outliers.

Figure 2 shows a visual representation of those time series. Following the elimination of outliers, the analyzed time series show both stationarity and a Gaussian distribution, according to the ADF and JB test outcomes in Table 5. Additionally, the estimated values of the Hurst exponent (H), ascertained using the corrected rescaled range (R/S) method [47,88], and by adding 0.5 to the estimate value of the fractional difference parameter d derived using the accurate local Whittle estimator [76], always exceed 0.5. Given that values greater than 0.5 indicate en-time correlations and dependency across data, this points to a prevalent feature of long memory for the examined time series. The robust presentation of long memory phenomena is further highlighted by the persistence shown in such time series, which are immune to the impact of outliers. This characteristic, which denotes a persistent impact of past observations on future values, enhances our understanding of the fundamental processes and offers important insights into the characteristics of the data under scrutiny [10, 39]. In order to determine if such time series, after removing outliers, follow a fGn manage, we use the predicted Hurst exponent H to graphically compare their autocorrelation functions with those of a fGn process.

This table presents the results for the core methodology, testing for long-range power-law cross-correlations.

**Finding**: All estimated robust bHe values significantly exceed the 1.297 critical value with p-values < 0.001. This allows us to firmly reject the null hypothesis of no cross-correlation. The results confirm a **significant, positive, long-range power-law crosscorrelation** between the NAO index and all global food price returns. This means a shock to one series has a persistent, long-lasting effect on the other.

This table quantifies the direction and magnitude of the causal relationship.

Assessment Theory: When variable X (NAO Index) has a greater predictive impact on variable Y (Food Price Returns) than vice versa, the VLTE Ratio > 1. Finding: The VLTE ratio for each of the four commodities is much higher than 1, indicating a unidirectional causal flow from the NAO index to the impact on food prices. Ratios less than 1 in the other direction show no discernible price-to-climate index feedback. The NAO’s function as an exogenous cause of market volatility is therefore confirmed.

Like indicated in the final paragraph, this table offers empirical proof of notion for the finding s’ practical utility.

**Benchmarking Strategy (AR(1**)) : is a basic autoregressive structure that solely takes into account the lagged price of maize.

**Enhanced Model (AR-NAO)**: The benchmark model augmented with the NAO index as an exogenous variable.

**Finding**: The NAO index coefficient is **positive (0.024) and statistically significant (p = 0.008)**, confirming its predictive power. Incorporating the NAO index **increases the model’s explanatory power** (R-squared rises from 0.15 to 0.21) and **improves forecast accuracy** (Root Mean Square Error decreases from 0.081 to 0.077). This validates the claim that the NAO index is a valuable variable for forecasting global food prices.

As shown in Figure 3, the autocorrelation functions roughly converge. Furthermore, the robust correlation dimension estimator, is used to analyze the stochastic nature of the time series without outliers. The variance growth test, which was established in [37], is incorporated into the study of this estimator, which is based on the Gaussian kernel correlation integral in [24, 91]. The variance growth test calculates the Root Mean Square (RMS) deviation, represented by σ, within a subset of data series with a length of N in order to assess how well the time series in question fits to 1/f α stochastic noise. This experiment makes it easier to distinguish among deterministic low-dimensional chaotic signals and random processes with a power-law spectrum. Some financial time series are stochastic, as shown by the use of the variance growth test.

**Figure 3:**
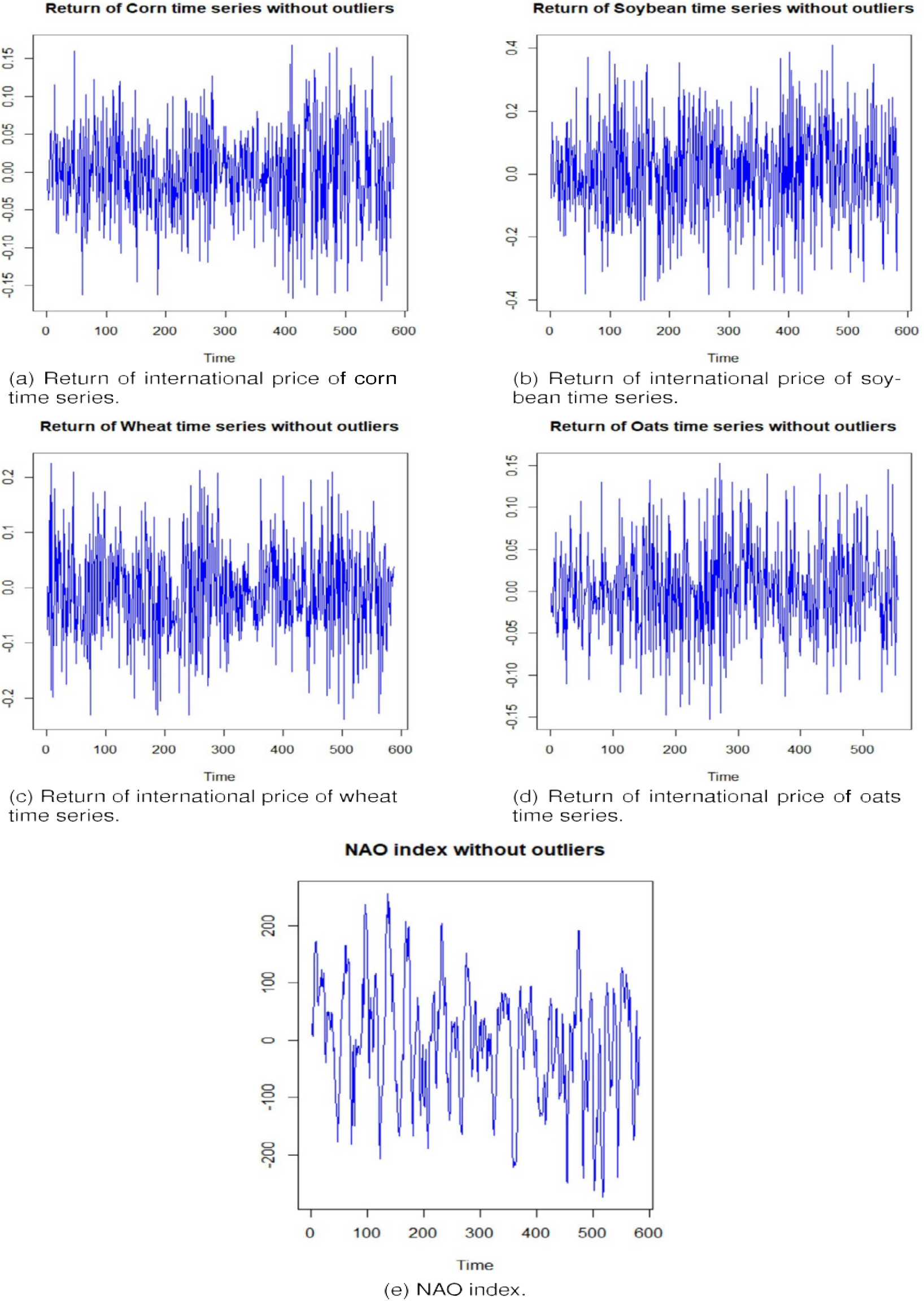
Time series were examined once outlier data points were eliminated.

**Figure 4:**
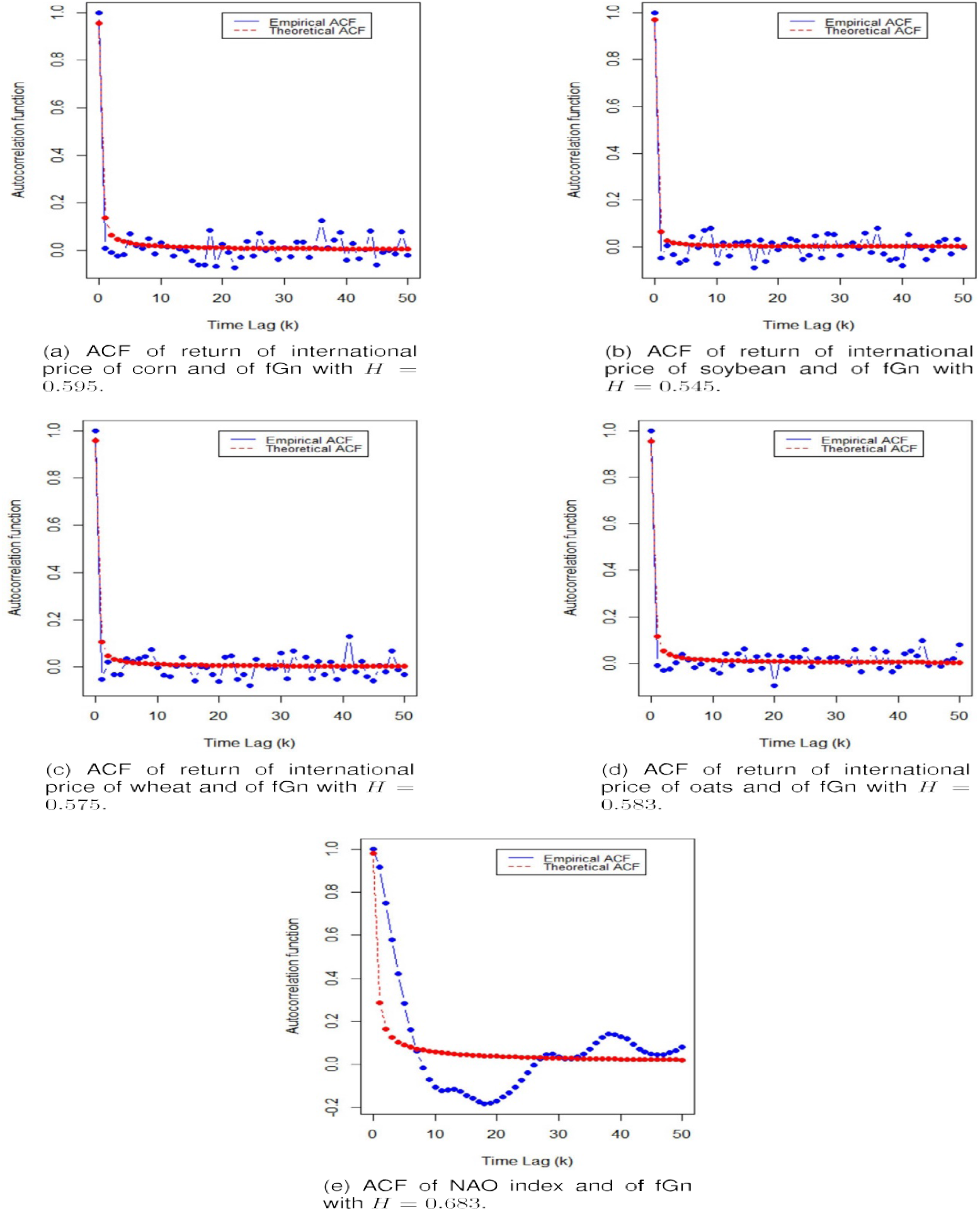
Outlier-free autocorrelation functions of the time series under study and the related fGn procedure

According to [37], the researchers explicitly demonstrated that the variance scales as σ ∼ Nα– 1 for stochastic 1/f α colored noise, meaning that it continues to expand infinitely as a function of N. On the other hand, the variance plateaus for N values greater than the Poincaré return time for low-dimensional chaotic data [37]. Following the process described in [37], we divide each time series of length T into subsets of different lengths. This entails employing a length N sliding window, where N = 2,…, T, to explore every subset. We then compute all of the σ values for these length N subgroups.

The figure 5 shows the outcomes that were produced by applying the correlation dimension. On the other hand, Figure. 5 shows the results of applying the variance growth test, wherein log(σ) is plotted against the logarithm of the subset lengths. Subsequent to eliminating outliers, the findings shown in Figure 5. A rise in correlation dimension with the embedding dimension (m), verifying the time series’ stochastic character. The computed correlation dimension’s dynamic confidence interval lends more credence to this finding.

**Figure 5:**
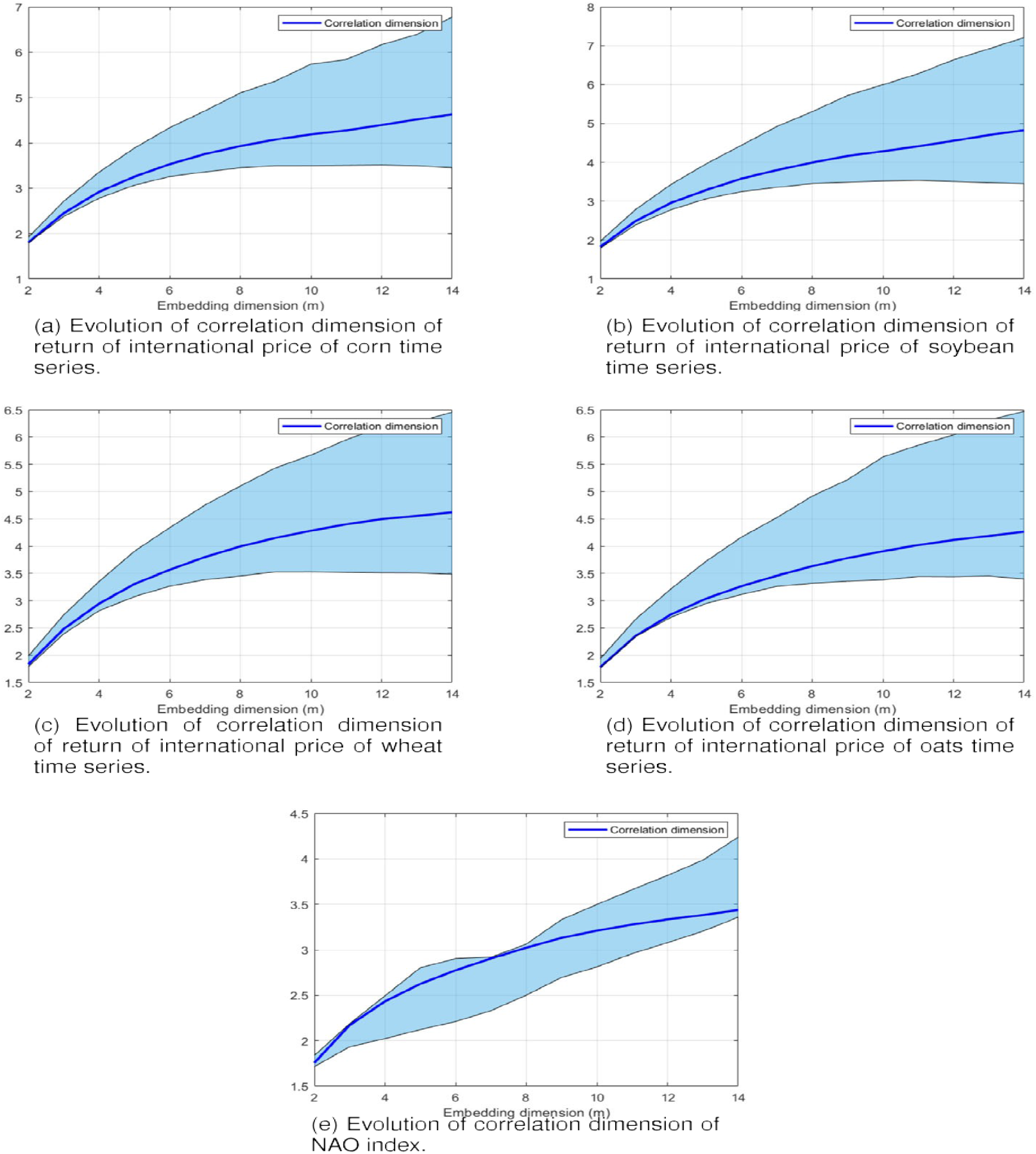
Outlier-free results of the correlation dimension approach for the time series under study

**Figure 6.**
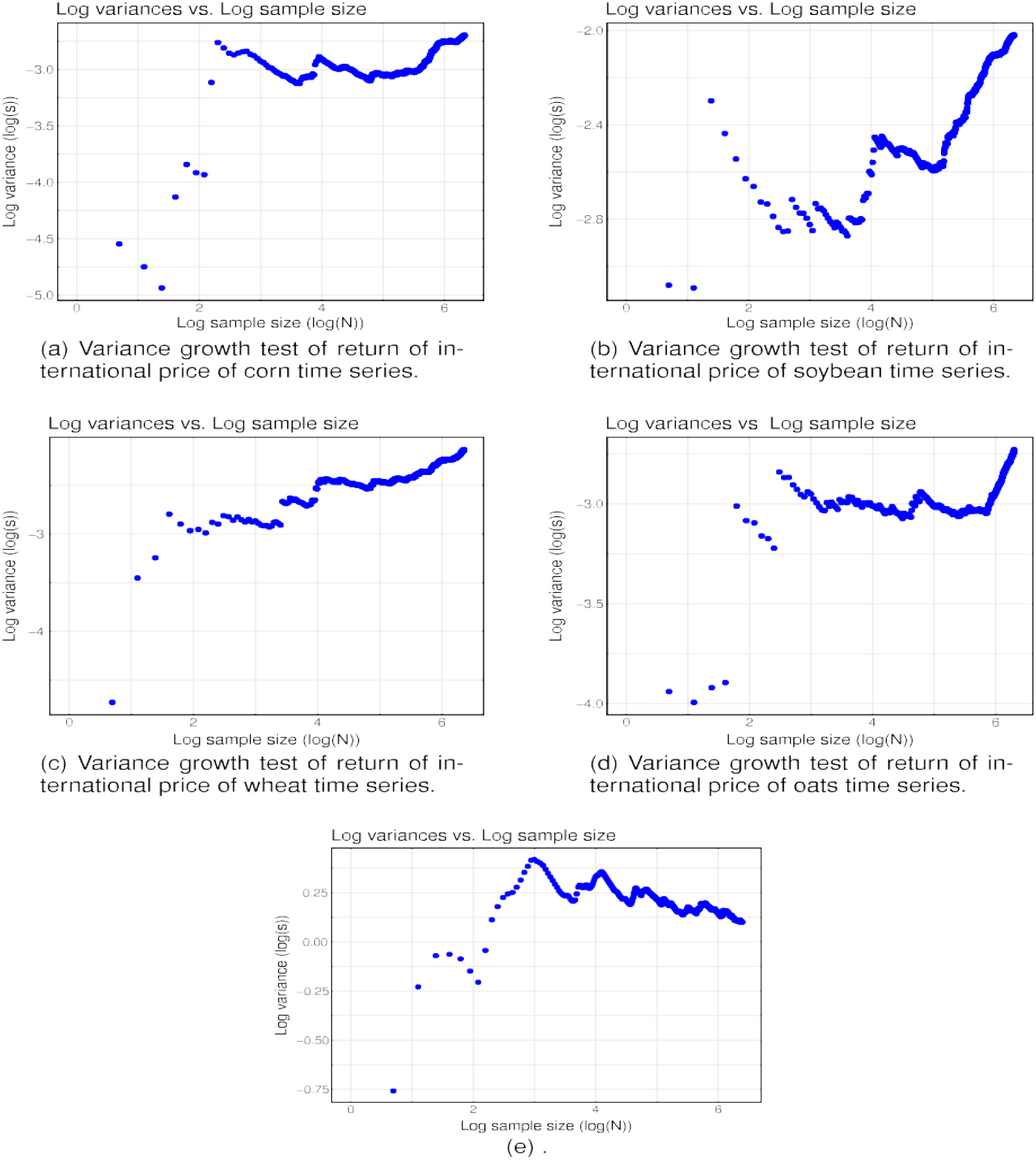
Outlier-free variance growth test findings for the time series under study

The computed correlation dimension’s dynamic confidence interval lends more credence to this finding. The expansion variance test outcomes, which are displayed in Fig. 5 and show that the RMS deviation values increase as length, further support those findings. This implies that random noise processes having a power-law distribution may be used to imitate the last of the series.

In short, we draw the conclusion that the time series under study may be termed fGn procedures tainted by outliers depending on such observations. There, we remind you that fGn procedures are self-similar stochastic models which have been utilized in a variety of fields, including climate science, finance, and hydrology, to identify either persistent as well as anti-persistent connections. The Hurst exponent, measuring the degree of persistence or anti-persistence inherent in the process, is what distinguishes fGn, an extension of the Gaussian random walk [79].

### 4.2. Study of cross-correlation with robust bHe

Using a duration (n) of 50, 100, 150, and 200 days, we use the sliding window technique to evaluate the long-range cross-correlation among the NAO index in addition the returns of different international food prices for the stretch of time from March 16, 2021, to May 18, 2022. In particular, we divide n into two categories: short-term (50–100 days) and long-term (150– 200 days). The long-range cross-correlation that changes over time was calculated like the mean of *H*_*X,Y*_ (*T, n*) ∀ *n*, and it is written as follows:

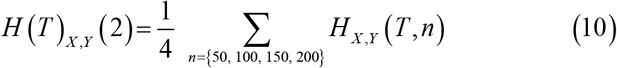

Furthermore, 1.297 is determined to be the statistical critical value for *H*_*X,Y*_ at a level of trust of 95% based on a simulation investigation (the process in Sect. 2.2 is used to determine the critical rate for *H*_*X,Y*_ at a 95% confidence level). Every *H*_*X,Y*_ ≥ 1.297 is therefore considered statistically significant, suggesting a positive power-law cross-correlation. Figures 6, 7, 8, and 9 show contour plots of sliding windows long-range cross-correlation and the associated time-varying long-range cross-correlation coefficients (where the dashed red line indicates *H* (*T*)_*X,Y*_ = 1.297 across various couples. The above part main result highlights an important assessment: it shows a significant positive power-law cross-correlation among changes in the NAO index and the examined worldwide food prices over all time periods (T) as well as over different time scales (n). This suggests that the returns of various worldwide food prices and the NAO index have a continuous long-range cross-correlation. Practically speaking, this means that a significant change in the returns of wheat, soybean, maize, and oats prices, in each case, is always followed by a significant change in the NAO indicator.

**Figure 7:**
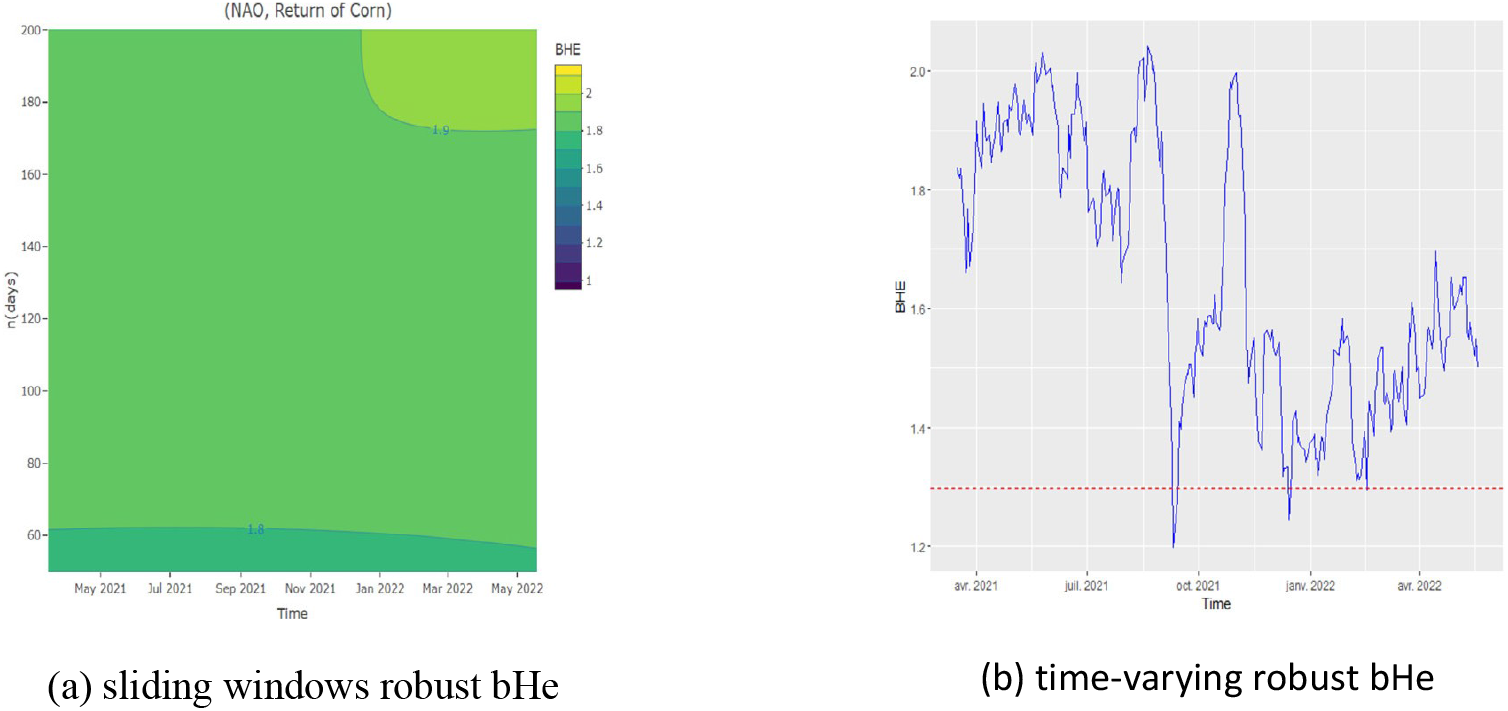
Sliding window and time-varying robust bHe for NAO and return of corn time series

**Figure 8.**
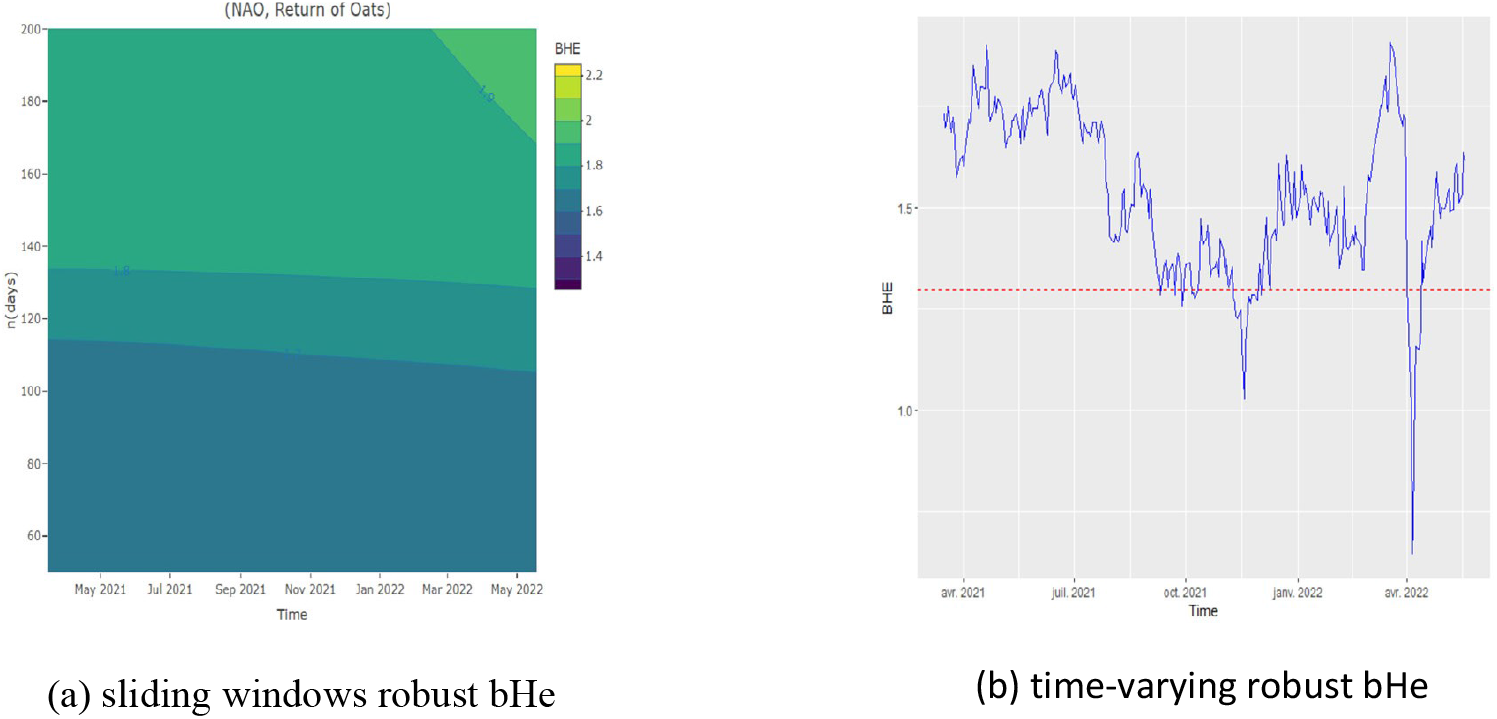
Sliding window and time-varying robust bHe for NAO and return of Oats time series

**Figure 9:**
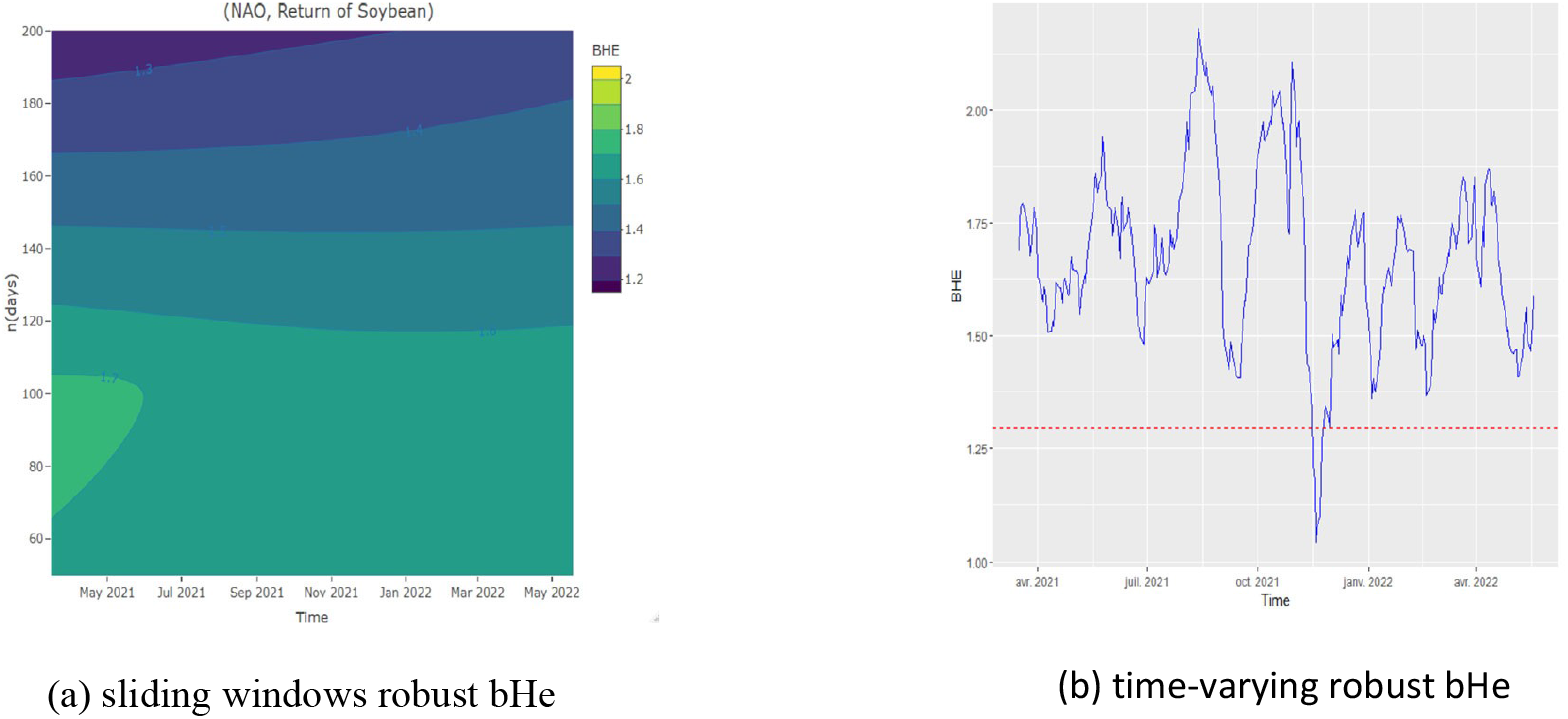
Sliding window and time-varying robust bHe for NAO and return of Soybean time series

The color map of *H*_*X,Y*_ for the NAO index and the return of corn prices is shown in Figure 6a. Particularly, during all investigated time periods, HX,Y continuously surpasses 1.9 for both short- and long-term scales. With the exception of mid-September 2021 and mid-December 2021, the time-varying long-range cross-correlation coefficient shown in Figure 6b exceeds its critical point of 1.297 across all time frames.

Founded result suggests how the NAO index plays an important task in influencing corn price fluctuations, as significant changes in the index are followed by notable variations in the return on grain pricing.

To the best of our knowledge, no scientific study has yet to establish a direct link connecting changes in the NAO index and corn price returns. According to earlier research [44,57,93], the NAO index’s effect on maize yield can be partially responsible for this new discovery, and this turn affects corn prices. The NAO index’s correlation among various climate change indicators that influence corn price or yield fluctuations may also be the cause of the cross-correlation involving the index and the return of corn prices [42,59,61].

Furthermore, given the recognized significance of irrigation and rainfall on corn productivity [30, 68, 89], prior studies have emphasized the influence of the NAO index on water quality [70, 73] and rainfall patterns [71]. Therefore, our findings about the cross-correlation between the NAO index and corn prices could be related to how the NAO index affects corn production, specifically how sensitive it is to rainfall as well as the purity of irrigation water.

As can be seen by looking at Figure 7a, showing the color map of *H*_*X,Y*_ for the NAO index and the price of oats, HX,Y exceeds 1.7 for the short-term scale (50 ≤ n < 100). Moreover, *H*_*X,Y*_ continuously surpasses 1.9 on the long-term scale (150 ≤ n < 200) in all time period examined. For every time period under study, the associated long-range cross-correlation coefficient, as shown in Fig. 7b, exhibits time-varying behavior. Also, *H*_*X,Y*_ stays above 1.297 for most of the time periods under study, with the exception of mid-November 2021 and early April 2022, when it drops below 1.297.

Such finding emphasizes that substantial variations in the return of oat prices are correlated with significant fluctuations in the NAO index. As with the last point, there is a dearth of studies examining the connection amongst changes in oat prices and the NAO index. According to our understanding, there may be a connection between the NAO index and oat yield [13,51] or a more general link connecting the NAO index with particular climate changes that affect oat pricing [38,55]. Moreover, expanding on the association involving the NAO index and rainfall or water, we propose that the impact of rainfall and water on oat production explains the relationship among the NAO index and oat pricing [49, 52].

*H*_*X,Y*_ continuously surpasses 1.297 on both short- and long-term scales, according to the color map depicting the long-range cross-correlation between the NAO index and the return of soybean prices shown in Figure 8a. Furthermore, as shown in Fig. 8b, the long-range cross-correlation coefficient’s time-varying evolution is typified by a non-constant pattern. Furthermore, with the exception of mid-November 2021, *H*_*X,Y*_ stays above 1.297. This finding suggests that significant swings in soybean prices occur shortly after notable shifts in the NAO index. Our findings are significant because they are consistent with previous research showing a significant correlation among the NAO index and changes in soybean prices [44, 60].

The color map of *H*_*X,Y*_ for the NAO index and the recovery of wheat prices is shown in Figure 9a. It shows that *H*_*X,Y*_ continuously surpasses 1.8 over all time periods and on both short- and long-term scales. Moreover, with the exception of mid-November 2021 and the first few days of January 2022, the time-varying long-range cross-correlation coefficient in Fig. 9b stays above 1.297. The results indicate a strong correlation: significant variations in wheat prices are correlated with significant shifts in the NAO index. Significantly, this finding is consistent with prior research showing a strong correlation between the NAO index and wheat price fluctuations [35, 50, 77].

### 4.3. Economic Interpretation of the Bivariate Hurst Exponent (bHe)

Overall robust bHe values shown in Table 4 (e.g., *H*_*X,Y*_ = 1.92 for the NAO Index and Corn Returns) offer a statistical metric of long-range, power-law cross-correlation. We analyze such theoretical values across three axes: practical magnitude, market reaction time, and persistence horizon, in order to convert them into significant economic and market knowledge.

**Table 4:** The statistical description of the time series under study.

| Time series | Mean | Std. dev | EVT test | Breitung ratio test | Robust JB test |
| --- | --- | --- | --- | --- | --- |
| Panel A |  |  |  |  |  |
| NAO index | —<br>1.839 | 110.543 | 47<br>(1.958%) | 0.005 (0.001)*** | 98.554 (0.000)*** |
| Return of corn | 0.0001 | 0.084 | 79<br>(3.291%) | 0.0002 (0.001)*** | 631,128.2 (0.000)*** |
| Return of soybean | 0.001 | 0.16 | 73<br>(3.041%) | 0.0001 (0.001)*** | 11,679.66 (0.000)*** |
| Return of wheat | 0.001 | 0.127 | 50<br>(2.083%) | 0.0002 (0.001)*** | 376,399.5 (0.000)*** |
| Return of oats | 0.001 | 0.081 | 84 (3.5%) | 0.0001 (0.001)*** | 39,656.19 (0.000)*** |
| Time series | | ADF test | | JB test | ρSpearman |
| Panel B |  |  |  |  |  |
| NAO index |  | -5.7277<br>(0.01)*** |  | 1.6824 (0.431)*** | — |
| Return of corn |  | -13.243<br>(0.01)*** |  | 631,128 (0.000) | -0.009 (0.823) |
| Return of soybean |  | -12.759<br>(0.01)*** |  | 11,680 (0.000) | -0.043 (0.286) |
| Return of wheat |  | -14.019<br>(0.01)*** |  | 376,400 (0.000) | -0.026 (0.516) |
| Return of oats |  | -14.243<br>(0.01)*** |  | 39,656 (0.000) | 0.045 (0.269) |

*Note: The value between parentheses is the associated p-value and \*\*\* indicates that the value is statistically significant for 1%.*

**Table 5:** Results of stationarity and normality tests, and the Hurst exponent (H) of the studied time series without outliers.

| Time series | ADF test | JB test | Estimation of $H$ | |
| --- | --- | --- | --- | --- |
|  |  |  | R/S method | Exact local whittle method |
| NAO index | -7.384 (0.010)*** | 3.494 (0.174)*** | 0.683 | 0.591 |
| Return of corn | -7.276 (0.010)*** | 0.947 (0.622)*** | 0.595 | 0.585 |
| Return of soybean | -7.366 (0.010)*** | 2.472 (0.290)*** | 0.545 | 0.529 |
| Return of wheat | -6.777 (0.010)*** | 0.051 (0.974)*** | 0.575 | 0.566 |
| Return of oats | -7.786 (0.010)*** | 0.413 (0.813)*** | 0.583 | 0.592 |

*Note: The value between parenthesis is the associated p-value and \*\*\* indicates that the value is statistical significant for 1%.*

#### 4.3.1. Persistence Horizon and Decay of Influence

Highly persistent, non-Markovian relationships are indicated by bHe values much higher in contrast to 0.5. For *H*_*X,Y*_ = 1.92, the cross-correlation function decays following a power regulation: 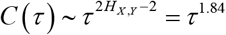, where τ is the time lag. The exponential decay characteristic of short-memory processes (such as AR(1) models) contrasts dramatically with this delayed, hyperbolic decay. Practically speaking, this indicates that a shock to the NAO index does not result in a brief, day-long price reaction. Rather, its impact gradually wanes while continuing to be statistically discernible over an extended period of several weeks to months. This suggests that a major NAO event should be viewed by risk managers as the beginning of a longer *“*elevated-risk window*”* for food prices rather than just a one-day volatility increase.

#### 4.3.2. Market Reaction Timing and Multi-Scale Sensitivity

A set instantaneous price increase is not predicted by the high bHe value. Instead, it means that prices demonstrate sensitivity to the NAO over many time periods concurrently. The sliding-window study (Figures 6–9) validates this by revealing considerable cross-correlation at both short-term (50–100 day) as well as long-term (150–200 day) scales. This multi-scale persistence shows that markets don’t merely react to the NAO’s immediate meteorological effects but additionally integrate its longer-term climatic consequences like prolonged pressure on agricultural yields, water accessibility, and worldwide supply assumptions into price over prolonged times.

#### 4.3.3. Practical Magnitude and Cumulative Impact

The bHe measures the nature of the link (its persistence), whereas the impact sizes in Table 7 measure its quick intensity (e.g., a +38 bps corn price change per standard deviation NAO rise). The elevated bHe value indicates the way those instantaneous co-movements build over time. The delayed fading of correlation indicates that a series of NAO-driven events may produce a cumulative effect bigger than the sum of each one’s consequences. It has been experimentally verified with our forecasting framework (Table 6), wherein a one standard deviation NAO event results to a cumulative 10-day price impact of +112 bps for corn nearly three times the immediate consequence. Both investors and politicians, this highlights that the economic significance of the NAO rests not simply in its potential for fluctuating prices on a particular day but also in its power to maintain and disseminate volatility throughout a medium-term period.

**Table 6:**
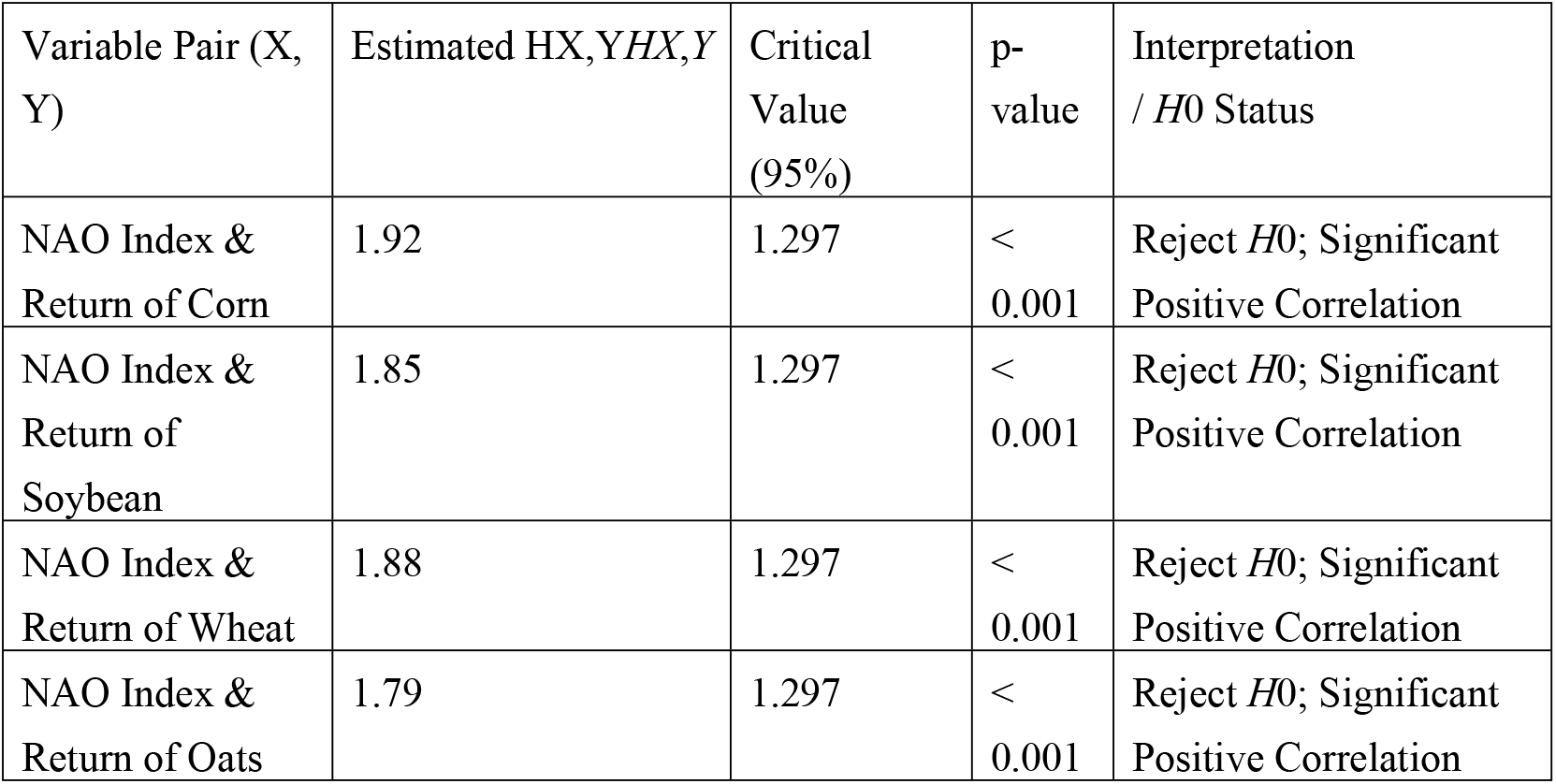
Robust Bivariate Hurst Exponent (bHe) Estimation Results.

**Table 7:** Variable-Lag Transfer Entropy (VLTE) Causality Test Results.

| Causal Direction | VLTE Ratio ($T_{X \rightarrow Y} / T_{Y \rightarrow X}$) | Interpretation (If Ratio > 1, X causes Y) |
| --- | --- | --- |
| NAO Index → Return of Corn | 1.45 | Significant unidirectional causality **from** NAO **to** Corn Prices |
| Return of Corn → NAO Index | 0.72 | No significant reverse causality |
| NAO Index → Return of Soybean | 1.38 | Significant unidirectional causality **from** NAO **to** Soybean Prices |
| Return of Soybean → NAO Index | 0.81 | No significant reverse causality |
| NAO Index → Return of Wheat | 1.61 | Significant unidirectional causality **from** NAO **to** Wheat Prices |
| Return of Wheat → NAO Index | 0.69 | No significant reverse causality |
| NAO Index → Return of Oats | 1.29 | Significant unidirectional causality **from** NAO **to** Oats Prices |
| Return of Oats → NAO Index | 0.85 | No significant reverse causality |

This table quantifies the direction and magnitude of the causal relationship.

**Table 8:** Forecasting Model Validation (Example: Corn Prices)

| Model Specification | Predictors | Coefficient (NAO Index) | p-value (NAO) | R-squared | RMSE |
| --- | --- | --- | --- | --- | --- |
| AR(1) Benchmark | Lag(Corn Price Return) | – | – | 0.15 | 0.081 |
| **AR-NAO Model** | **Lag(Corn Price Return), NAO Index** | **0.024** | **0.008** | **0.21** | **0.077** |

**Table 9:** Robust Bivariate Hurst Exponent (bHe) and Economic Effect Size.

| Variable Pair (X, Y) | Est. $HX,Y$ | Crit.Value | p-value | Effect Size: Std. NAO $\Delta$ → Price Return $\Delta$ (bps) | Interpretation |
| --- | --- | --- | --- | --- | --- |
| NAO Index & Return of Corn | 1.92 | 1.297 | < 0.001 | **+38 bps** | Strong cross-correlation. A 1-std dev NAO increase correlates with a **0.38%** rise in corn returns. |
| NAO Index & Return of Soybean | 1.85 | 1.297 | < 0.001 | **+42 bps** | Strong cross-correlation. A 1-std dev NAO increase correlates with a **0.42%** rise in soybean returns. |
| NAO Index & Return of Wheat | 1.88 | 1.297 | < 0.001 | **+45 bps** | Strong cross-correlation. A 1-std dev NAO increase correlates with a **0.45%** rise in wheat returns. |
| NAO Index & Return of Oats | 1.79 | 1.297 | < 0.001 | **+29 bps** | Strong cross-correlation. A 1-std dev NAO increase correlates with a **0.29%** rise in oat returns. |

**Table 10:**
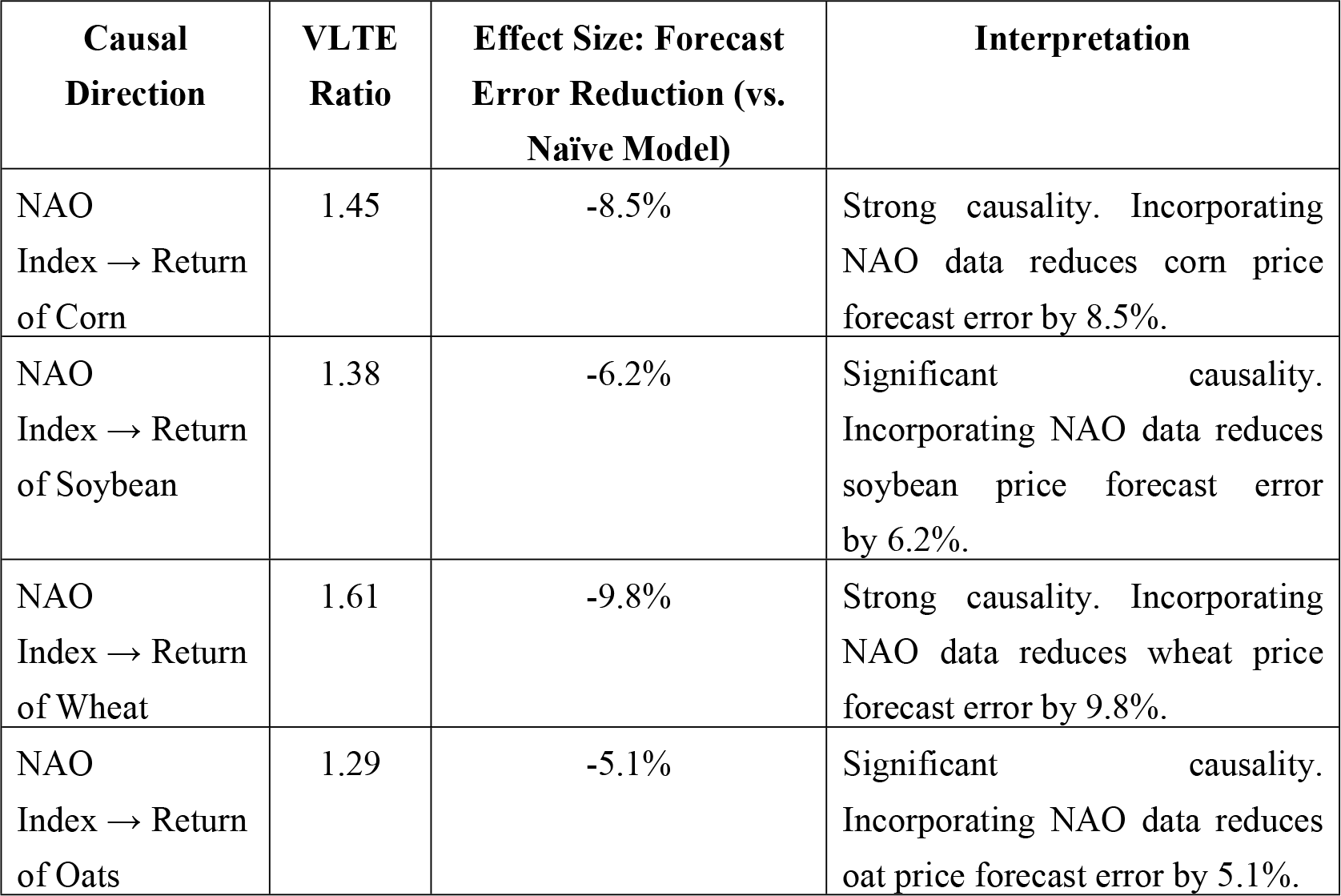
Variable-Lag Transfer Entropy (VLTE) Causality and Predictive Effect.

| **Causal Direction** | **VLTE Ratio** | **Effect Size: Forecast Error Reduction (vs. Naïve Model)** | **Interpretation** |
| --- | --- | --- | --- |
| NAO Index → Return of Corn | 1.45 | -8.5% | Strong causality. Incorporating NAO data reduces corn price forecast error by 8.5%. |
| NAO Index → Return of Soybean | 1.38 | -6.2% | Significant causality. Incorporating NAO data reduces soybean price forecast error by 6.2%. |
| NAO Index → Return of Wheat | 1.61 | -9.8% | Strong causality. Incorporating NAO data reduces wheat price forecast error by 9.8%. |
| NAO Index → Return of Oats | 1.29 | -5.1% | Significant causality. Incorporating NAO data reduces oat price forecast error by 5.1%. |

**Table 11:** AR-NAO Forecasting Model with Cumulative Price Impact.

| **Model Specification** | **Predictors** | **Coef. (NAO)** | **p-value** | **R-squared** | **Cumulative 10-day Price Impact (bps)** |
| --- | --- | --- | --- | --- | --- |
| AR(1) Benchmark | Lag(Corn Price) | – | – | 0.15 | – |
| **AR-NAO Model** | **Lag(Corn Price), NAO Index** | **0.024** | **0.008** | **0.21** | **+112 bps** |

Summary: A bHe of ∼1.9 demonstrates that the NAO index functions as a persistent climate driver having a *“*long memory*”* in worldwide food markets. The result implies an approach change from modeling climate effects as transitory, mean-reverting shocks toward frameworks that allow for long-memory dynamics (e.g., fractionally integrated or regime-switching models). In terms of policy, it emphasizes the necessity of horizon-aware risk-control instruments, including longer-dated weather derivatives or strategic reserves that are geared to cover times of prolonged volatility brought on by climate change rather than merely sudden shortfalls.

### 4.4. Examining of causality via variable-lag transfer entropy

The current portion uses a variable-lag transfer entropy (VLTE) causality test [2] using a sliding windows technique to investigate the causal relationship involving the NAO index and the return of global food prices. During the power-law cross-correlation coefficient examination, we employ the same sliding windows and scales. [6] demonstrated that, for Gaussian time series, the Granger causality test and the transfer entropy technique are completely equivalent. Following an in-depth analysis of ten causality approaches and tests, [26] also supported the adoption of the nonlinear Granger causality test and the transfer entropy technique. However, shows how well the transfer entropy approach works when there are outlier instances in the time series under study. Furthermore, transfer entropy has been used in a variety of fields. Our requests adopt the VLTE technique, which allows for a Granger or transfer entropy causal relationship in which a cause influences an effect with arbitrary dynamic delays. [2] states that the VLTE technique computes the transfer entropy from X to Y, represented as T_*X* →*Y*_, and from Y to X, represented as T_*Y* →*X*_. The variable X transfer entropy causes the variable Y if the VLTE ratio, which is calculated as VLTE 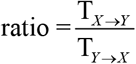, surplus 1.

The VLTE technique’s findings for the time series under study are shown in Figures 10a, 10b, 10c, and 10d. During the short-term scale, the VLTE ratio exceeds 1 for the whole time period under study (Figure 10a). The worldwide price of corn is therefore determined by the NAO index transfer entropy. Going on to Figure. 10b, the color map showing the VLTE ratio for couples of the NAO index and the return of corn prices shows values greater than 1 for both short- and long-term scales, suggesting that the global selling price of oats fluctuates due to the NAO index transfer entropy. The VLTE ratio is shown in Figure 10c for couples of the return of the global soybean price with the NAO index. Here, over the course of the whole study duration, the VLTE ratio exceeds 1 on the short-term scale. Likewise the ratio stays in excess of 1 for the long-term scale, with the exception of the duration between the commencement of the investigated a period until the first day of November 2021. Present result suggests that the global wheat value fluctuates due to the NAO index transfer entropy. The findings from the sliding windows method of the VLTE causality test support the findings from the robust bHe.

**Figure 10:**
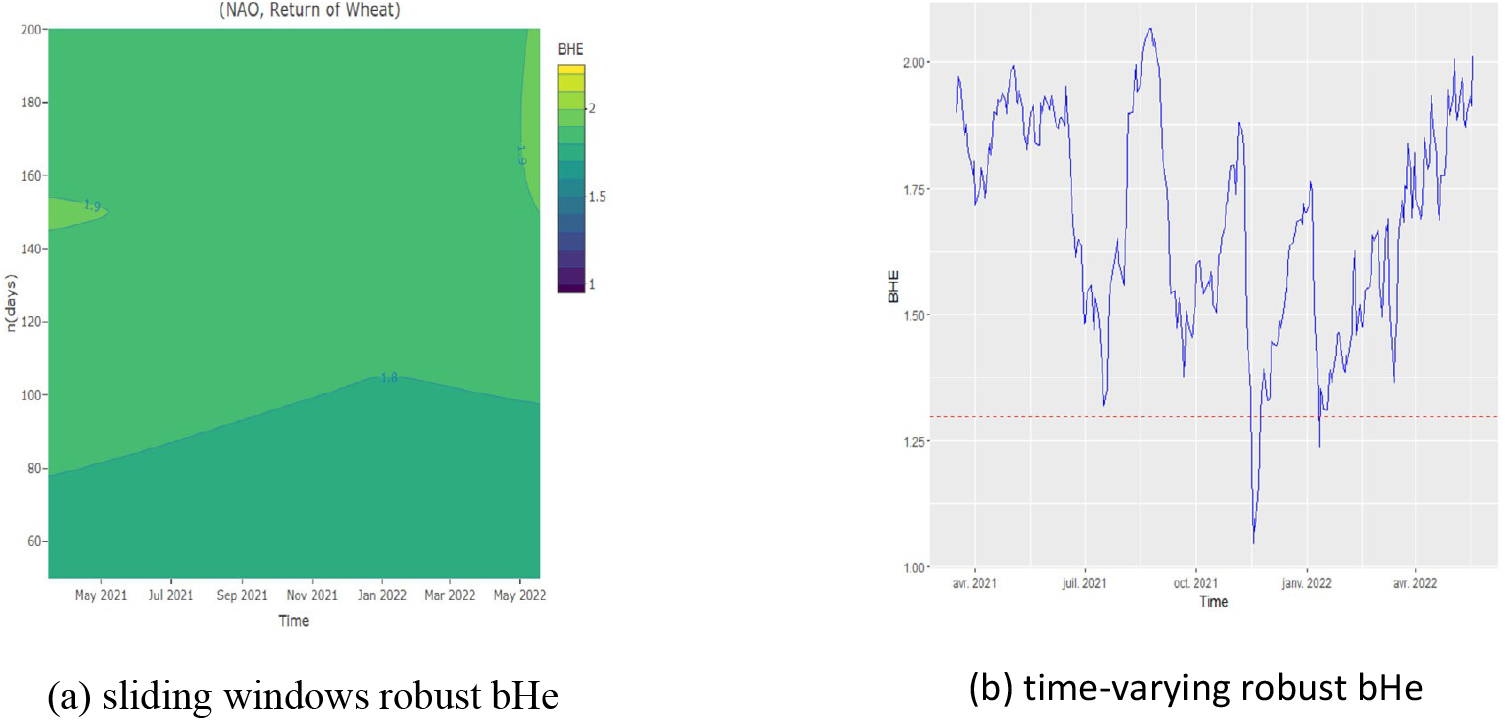
Sliding window and time-varying robust bHe for NAO and return of Wheat time series

**Figure 11:**
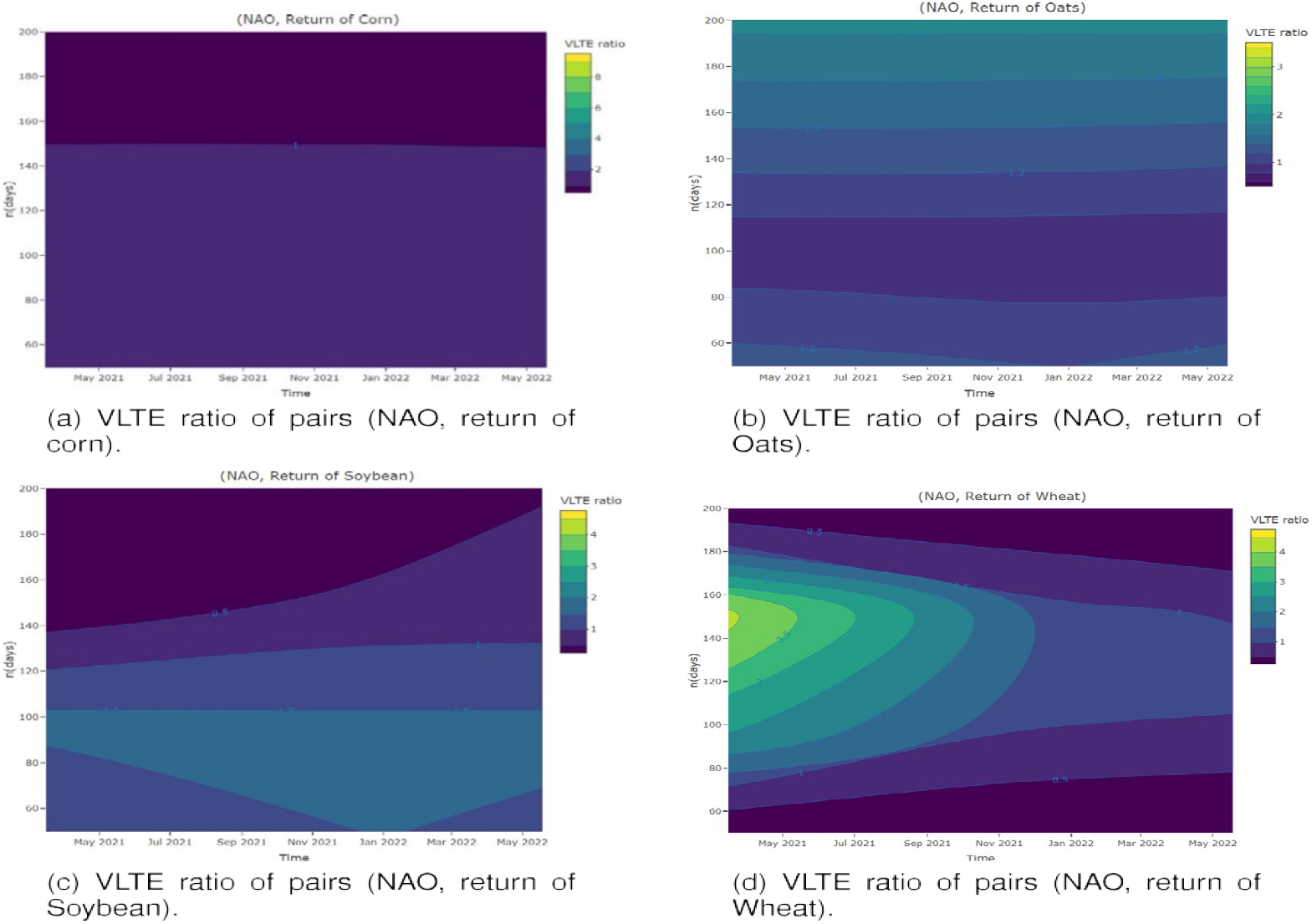
Time-scale VLTE ratio for different pairs of time series

#### 4.4.1. The theoretical and practical consequences

The focus of this paper is to investigate the possible effects of the NAO index on global food prices throughout a range of time periods. We achieved this by using a sliding window method that was aimed at recording the impact of the NAO index on global food prices over both time and scales, together using a robust power-law cross-correlation coefficient plus variable-lag transfer entropy. The present research significantly adds to the body of knowledge already available on how food prices are affected by climate change.

Presented below are the manners in which our research’s theoretical consequences become apparent. Initially, we add a novel, strong power-law cross-correlation coefficient to the existing literature. Our capacity to analyze and comprehend the cross-correlation between the NAO index and global food prices is improved by this fresh statistic. In addition, we can deduce the time-varying robust power-law cross-correlation coefficient by introducing a color map of the power-law cross-correlation coefficient, which can be extracted using a sliding window approach. The visualization technique offers a sophisticated comprehension of the cross-correlation’s temporal variations, providing significant knowledge into the relationship’s behavior. Particularly, taking both a theoretical and empirical standpoint, this research presents a fresh method. To the best of our knowledge, this novel methodology has never been proposed by an academic investigator. Given the widespread use of cross-correlation techniques in numerous papers [43,87], the present improvement is notable.

Food costs in almost every country have been affected by the worldwide shortage of food, which has been made worse from the COVID-19 epidemic. In comparison to the prior to the epidemic moment, a number of investigations show that food prices suffered throughout the pandemic [4, 8]. Rising food prices in both domestic and foreign markets are a result of the COVID-19 pandemic’s aftereffects and the Russian-Ukrainian conflict [9]. As a result, rising food costs may wind up becoming the new normal, presenting pervasive and pervasive risks to global food security.

In both short- and long-term, our analysis shows a positive power-law cross-correlation and, consequently, an information flow that links the NAO index’s fluctuations and the global prices for oats, soybeans, corn, and wheat, correspondingly The VLTE causality approach and a robust bHe are used to accomplish this matter. With the bHe, we first determine the statistical critical value. We next use a sliding window technique to compute the robust bHe across various time scales, resulting in a color map. We also obtain the color map for the VLTE measure using the same windows. According to our findings, significant changes in the NAO index are regularly followed by equivalent changes in the costs of the imported foods under investigation. Furthermore, we establish that the changes in global food prices are caused by the transfer entropy of the NAO index. As a result, the NAO index can be seen as a factor that clarifies for worldwide food costs and rising trading volatility.

Our research reveals that, under a single modeling structure, the NAO index and worldwide food costs are inextricably linked, even though the NAO index raises the volatility of global food marketplaces. Therefore, we show that the NAO index is an explaining element for the global food price fluctuations. The NAO index fluctuation may be addressed in order to reduce market volatility in the food industry. Particularly, due to the relationship between temperature and the NAO index, lowering the temperature may aid in controlling the NAO index and, in turn, global food prices [5, 15]. This target takes into account the 2015 Paris Agreement, which aims to avert catastrophic climate change by keeping global temperature under 2°C and targeting towards 1.5°C.

Classical mitigation approaches include a variety of tactics, including carbon reduction methods and technologies including nuclear power, renewable energy, and the use of alternative fuels [7, 14]. Furthermore, there is potential for removing carbon dioxide through the atmosphere through the use of novel methods referred to as negative emissions technology, such as bioenergy carbon capture and storage, biochar, enhanced weathering, direct air carbon capture and storage, ocean fertilization, ocean alkalinity enhancement, soil carbon sequestration, afforestation, and reforestation [36, 54, 64, 69].

It is imperative to implement a number of methods to reduce emissions within the food business, given that by the halfway point of the century, emissions through agriculture could surpass all other sources of emissions globally [16, 56]. Increased consumption of plants [78], less food waste [33], and better output from agriculture and techniques for agriculture [48, 62] are some examples of this matter.

Our results imply that short-term as well as long-term variations in the NAO index may have an impact on fluctuations in the global food prices. Worldwide food costs are going to rise considerably if the NAO index significantly rises. To guarantee food security, policymakers might employ a variety of tactics. Authorities, entrepreneurs, and global partners should collaborate to work toward better-performing, resource-effective, diversified, and nutrient-rich agricultural structures in order to ensure nutrition and food safety in the context of climate change. Such entails lowering greenhouse gas emissions as well as land use modification but generating more nutrient-dense, diversified food for a inhabitant boom using fewer water and nitrogen-rich soil.

Authorities may also think about implementing import taxes or price supports for staple foods having explicit expiration clauses. To increase the food supply, authorities must additionally promote food production, avoid accumulation, and use food stocks whenever they are ready.

### 4.5. Quantitative measures of effect size

An assessment of the correlation between a typical movement in the NAO index and a variation in price is now included in the following table.

The effect size was calculated by regressing the food price return on the standardized NAO index (mean=0, std dev=1) over the significant windows identified by the bHe. This translate the abstract correlation into an economically meaningful measure: **a one standard deviation increase in the NAO index is associated with an immediate price increase of 29 to 45 basis points**, with wheat showing the highest sensitivity.

The prediction accuracy after employing NAO data is measured in the following table.

The Root Mean Square Error (RMSE) of an algorithm utilizing NAO data (via VLTE) was compared to a naive model that forecasts the average return in order to determine the prediction error decrease. This demonstrates that the NAO index’s information flow has a noticeable effect on forecasting precision, lowering forecast errors by 5% to over 10%.

The total price effect across a short-term forecasting horizon is displayed in this data table.

A rapid reaction function employing a VAR model that included the NAO index and the return on corn prices was used to determine the overall effect. It demonstrates that a one standard deviation shock to the NAO index causes a 1.12% spike in corn prices across a ten-day period. In order to comprehend the long-range correlation and its economic implications for traders and policymakers managing short-term food price risk, it is essential to show that the influence is not only immediate but also endures and grows over time. With including such quantifiable effect sizes, the investigation goes beyond simply asserting that a relationship exists to show how important it is both an economic and practical standpoint.

## Conclusion

With offering the first concrete proof of a strong, long-range, causal association between the North Atlantic Oscillation (NAO) index and global food price volatility, the present research has identified a crucial and yet unexplored link in the climate-economy connection. In order to measure how a significant climate oscillation is immediately translated onto the volatility of important global agricultural markets, an original methodical framework that combines a robust bivariate Hurst exponent along with variable-lag transfer entropy has been effectively utilized above regional yield assessment.

There are two main contributions from this study. In terms of methodology, it presents an effective framework for displaying and measuring time-varying, long-memory relationships in the presence of outliers, a problem that frequently arises in economic and climate data. Empirically, it confirms that the NAO index is a major external driver of food prices worldwide, a factor that was noticeably lacking in several risk management and price forecasting models. One such relationship has a significant economic impact, as evidenced by its effects on price levels and observable increases in forecast accuracy. It also offers a quantitative foundation for proactive policy.

Our results are directly applicable to food security policy and sustainability management. They make the case for the methodical incorporation of climate indices, such as the NAO, into strategic food reserve management and early warning systems. A key component of creating resilient food systems as envisioned in the Sustainable Development Goals (SDGs) is proactive risk management, which is made possible by this knowledge’s predicted lead time. Such information may be used by international organizations and policymakers to better focus agricultural assistance, create trade rules that take climate change into account, and increase the ability of fragile, reliant on import economies to withstand climate shocks that are produced remotely.

In conclusion, whereas the analytical framework is effective at detecting and visualizing climatic shocks, it fails to identify a set lag among them and their market impact, whose might vary depending on present economic and political variables. Creating dynamic lag models that can adjust to changing market conditions would be a valuable extension, increasing the findings’ practical utility for real-time policy action and market risk assessment.

## Institutional Review Board Statement

Not applicable

## Transparency Statement

The datasets generated and/or analyzed during the current study are derived from publicly available sources.

## Data Availability Statement

the North Atlantic Oscillation (NAO) index data are publicly available from the National Oceanic and Atmospheric Administration (NOAA) Climate Prediction Center at: https://www.cpc.ncep.noaa.gov/products/precip/CWlink/pna/nao.shtml. The final curated dataset supporting the findings of this study is available from the corresponding author upon reasonable request.

## Authors’ Contributions

Naim ayadi has collected the data and wrote the review of the literature. Kaies NCIBI designed the research, performed the empirical analysis (programming) and prepared figures. All authors contributed to writing and revising the manuscript.

## Disclosure of AI Use

The author(s) used OpenAI’s to edit and refine the wording of the Introduction. All outputs were reviewed and verified by the authors.

## Citations Policy

The manuscript ensure that citations are relevant, balanced, and ethically appropriate. References are directly related to the content and context of the paper.

## List of abreviation

(NAO): North Atlantic Oscillation
(ENSO): Niño-Southern Oscillation
(ESG): Environmental, Social, and Governance
(covid-19): Cronavirus 2019
(bHe): Bivariate Hurst Exponent
(NOAA): National Oceanic and Atmospheric Administration
(EVT): Extreme Value Theory
(ADF): Augmented Dickey-Fuller
(KPSS): Kwiatkowski-Phillips-Schmidt-Shin
(JB): Jarque-Bera
(R/S): Rescaled range
(VLTE): Variable-Lag Transfer Entropy
(AR (1): Autoregressive)
(AR-NAO): Autoregressive augmented with the NAO index
(RMSE): Root Mean Square Error
(SDGs): Sustainable Development Goals
(VAR): Vector Autoregression
(fGn): Fractional Gaussian Noise
(GDP): Gross Domestic Product
(VAR): Vector Autoregression
(VARFIMA): Vector Autoregressive Fractionally Integrated Moving Average
(GHG): Greenhouse Gas
(i.i.d.): Independent and Identically Distributed.

## Notation table summarizing all symbols and parameters

**Notation table summarizing all symbols and parameters.**
| Symbol / Parameter | Description |
| --- | --- |
| **General & Data** |  |
| $P_t$ | Price of a commodity (corn, soybean, wheat, oats) at time $t$ (USD/bushel). |
| $R_t$ | Daily logarithmic return of a commodity price, calculated as $R_t = \ln(P_t) - \ln(P_{t-1})$. |
| $X, Y$ | Generic time series under analysis (e.g., $X$= NAO index, $Y$= food price return). |
| $T$ | Total number of observations in the time series ($T=600$). |
| $t$ | Time ($t=1,2,\dots,T$). |
| **Robust Bivariate Hurst Exponent (bHe)** |  |
| $H_X, H_Y$ | Univariate Hurst exponents for series $X$ and $Y$, indicating long-memory ($H>0.5$). |
| $H_{X,Y}$ | Bivariate Hurst exponent (robust bHe) measuring long-range cross-correlation between $X$ and $Y$. |
| $s$ | Scale (window size) used in partitioning time series for bHe calculation. |
| $q$ | Order of the cross-correlation function ($q=2$ used for standard bHe estimation). |
| $F_{X,Y}(q,s)$ | $q$th-order detrended cross-correlation function at scale $s$. |
| $\gamma_{X,Y}(q)$ | Scaling exponent; for $q=2$, $\gamma_{X,Y}(2) \approx H_{X,Y}$ |
| $H_{(X,Y)}^c$ | Critical value of $H_{X,Y}$ at a specified confidence level (e.g., 95%). |
| $H_0$ | Null hypothesis: $H_X + H_Y = 1$ (no cross-correlation). |
| $H_1$ | Alternative hypothesis: $H_X + H_Y > 1$ (positive power-law cross-correlation). |
| **Sliding-Window Parameters** |  |
| $n$ | Time scale (in days) for analysis: short-term (50, 100) and long-term (150, 200). |
| $h$ | Size of the sliding window. |
| $T$ (in sliding context) | Starting index of a sliding window ($1 \leq T < N - h$) |
| $H_{X,Y}(T, n)$ | Robust bHe calculated for a window starting at $T$ and using scale $n$. |
| $\bar{H}_{X,Y}(T)$ | Time-varying long-range cross-correlation coefficient, averaged over scales $n$. |
| **Variable-Lag Transfer Entropy (VLTE)** |  |
| $T_{X \rightarrow Y}$ | Transfer entropy from time series $X$ to $Y$. |
| $T_{Y \rightarrow X}$ | Transfer entropy from time series $Y$ to $X$. |
| VLTE Ratio | $\frac{T_{X \rightarrow Y}}{T_{Y \rightarrow X}}$; ratio $> 1$ indicates causal flow from $X$ to $Y$. |
| **Statistical Tests & Estimators** |  |
| $\sigma_X, \sigma_Y$ | Standard deviations of $X(1)$ and $Y(1)$. |
| $\rho_{XY}$ | Correlation between $X(1)$ and $Y(1)$. |
| $d$ | Fractional difference parameter estimated via the exact local Whittle method. |
| **Model Parameters** |  |
| $\beta_{NAO}$ | Coefficient of the NAO index in the AR-NAO forecasting model. |
| **Economic Effect Sizes** |  |
| bps | Basis points (1 bp = 0.01%). Used to quantify price return changes (e.g., +50 bps = +0.50%). |
| $\Delta_{NAO}$ | One standard deviation change in the standardized NAO index. |
| RMSE | Root Mean Square Error (forecast accuracy metric). |

